# Fidelity and adherence to a liquefied petroleum gas stove and fuel intervention: the multi-country Household Air Pollution Intervention Network (HAPIN) trial

**DOI:** 10.1101/2023.06.20.23291670

**Authors:** Kendra N. Williams, Ashlinn Quinn, Hayley North, Jiantong Wang, Ajay Pillarisetti, Lisa M. Thompson, Anaité Díaz-Artiga, Kalpana Balakrishnan, Gurusamy Thangavel, Ghislaine Rosa, Florien Ndagijimana, Lindsay J. Underhill, Miles A. Kirby, Elisa Puzzolo, Shakir Hossen, Lance A. Waller, Jennifer L. Peel, Joshua P. Rosenthal, Thomas F. Clasen, Steven A. Harvey, William Checkley, the HAPIN Investigators

## Abstract

**Background:** Reducing household air pollution (HAP) to levels associated with health benefits requires nearly exclusive use of clean cooking fuels and abandonment of traditional biomass fuels.

**Methods:** The Household Air Pollution Intervention Network (HAPIN) trial randomized 3,195 pregnant women in Guatemala, India, Peru, and Rwanda to receive a liquefied petroleum gas (LPG) stove intervention (n=1,590), with controls expected to continue cooking with biomass fuels (n=1,605). We assessed fidelity to intervention implementation and participant adherence to the intervention starting in pregnancy through the infant’s first birthday using fuel delivery and repair records, surveys, observations, and temperature-logging stove use monitors (SUMs).

**Results:** Fidelity and adherence to the HAPIN intervention were high. Median time required to refill LPG cylinders was 1 day (interquartile range 0-2). Although 26% (n=410) of intervention participants reported running out of LPG at some point, the number of times was low (median: 1 day [Q1, Q3: 1, 2]) and mostly limited to the first four months of the COVID-19 pandemic. Most repairs were completed on the same day as problems were reported. Traditional stove use was observed in only 3% of observation visits, and 89% of these observations were followed up with behavioral reinforcement. According to SUMs data, intervention households used their traditional stove a median of 0.4% of all monitored days, and 81% used the traditional stove <1 day per month. Traditional stove use was slightly higher post-COVID-19 (detected on a median [Q1, Q3] of 0.0% [0.0%, 3.4%] of days) than pre-COVID-19 (0.0% [0.0%, 1.6%] of days). There was no significant difference in intervention adherence pre– and post-birth.

**Conclusion:** Free stoves and an unlimited supply of LPG fuel delivered to participating homes combined with timely repairs, behavioral messaging, and comprehensive stove use monitoring contributed to high intervention fidelity and near-exclusive LPG use within the HAPIN trial.

## INTRODUCTION

Household air pollution (HAP), caused by the inefficient combustion of polluting fuels such as wood, dung, coal, charcoal, crop residue, and kerosene for household energy needs, was responsible for an estimated 3.2 million premature deaths and 82 million healthy life years lost in 2019.^1^ Transitioning to energy sources that are cleaner at the point of use, such as liquefied petroleum gas (LPG), electricity, natural gas, or ethanol, can significantly reduce exposure to HAP.^2, 3^ Exposure to HAP is associated with an array of poor health outcomes, including diabetes, kidney diseases, chronic respiratory diseases, cardiovascular diseases, maternal and neonatal disorders, respiratory infections, and tuberculosis.^4, 5^ However, there is a lack of evidence from randomized controlled trials on whether switching to cleaner fuels improves health.

Although some stove intervention trials have found substantial exposure-response relationships between specific air pollutants and health outcomes^6, 7^, most have not demonstrated significant health impacts of the interventions in intention-to-treat analyses.^8–11^ These null findings, however, have largely been ascribed to a lack of exclusive use of the clean fuel stove and HAP exposure levels that remained above both interim target and guideline levels of PM_2.5_ recommended by the World Health Organization (WHO).^12^ Recently, a small (n=180) randomized controlled trial of an LPG intervention in Peru was able to achieve near exclusive LPG stove use (>98% of total cooking minutes across all stoves in the household were done with LPG) and an average personal exposure to PM_2.5_ below the WHO interim target 1 in intervention participants (mean of 30 μg/m^3^ compared to 98 μg/m^3^ in control participants). However, the study found no impact of the stove intervention on blood pressure, lung function, or respiratory symptoms among non-pregnant adult women.^13^

The Household Air Pollution Intervention Network (HAPIN) randomized controlled trial sought to assess the effects of an LPG stove and fuel intervention in populations relying chiefly on solid biomass fuels (wood, charcoal, dung, agricultural residue) on four primary outcomes: infant birth weight, stunting at one year of age, and severe pneumonia in children under one year of age, and blood pressure in non-pregnant adult women living in the same household.^14^ The research questions raised by the trial required high levels of intervention fidelity and adherence in order to reach the levels of HAP believed necessary to achieve health benefits.^15^ Stoves and an unlimited supply of LPG were accompanied by behavior change approaches to attain as close to exclusive LPG use as possible.^16^

We previously reported high levels of intervention fidelity and adherence during pregnancy in the HAPIN trial as part of an assessment of pregnancy and birth outcomes.^17^ Here, we report on fidelity and adherence during the child’s first year of life and over the entire trial period, from LPG stove delivery through the child’s first birthday, as well as adherence in the subset of households in which a second non-pregnant adult woman was enrolled (to understand adherence in relation to the blood pressure primary outcome). We also explore the impact of the COVID-19 pandemic on intervention fidelity and adherence.

## METHODS

### Study Setting and Design

Details on the HAPIN study design have been published separately.^14, 16^ Briefly, we enrolled approximately 800 pregnant women between 9 to 19 weeks of gestation who primarily used biomass fuels for cooking in each of four countries: Jalapa, Guatemala; Puno, Peru; Kayonza, Rwanda; and Tamil Nadu, India. Participants who were randomly assigned to the intervention arm received an LPG stove, free LPG replacement cylinders delivered as needed, behavioral reinforcement to promote exclusive LPG use, and free stove repairs, if required. Control participants were expected to continue relying primarily on biomass cooking fuels. We also enrolled a subset of non-pregnant adult women (aged 40-79 years) living in the same household as the pregnant woman (n=417).

### Intervention and Delivery

Intervention participants received an LPG stove with at least two burners (see Quinn et al., 2021 for details on stove models) together with a continuous supply of free LPG fuel from two exchangeable cylinders.^17^ Households were instructed to request an LPG refill from the HAPIN study team when the first cylinder in their household ran out, thus, under typical consumption patterns, enabling them to continue cooking using their second cylinder and allowing HAPIN staff up to seven days to complete the refill request. At every LPG delivery visit in Guatemala, Peru, and Rwanda, and at every SUMs download visit in India, HAPIN field staff checked the functionality of the LPG stove and cylinders and recorded their observations in a survey. If a problem was identified, field staff either fixed it immediately or returned as soon as possible to resolve the issue. In between visits, participants were instructed to inform study staff if they had any problems with the LPG equipment. Dates of repair requests and repair completion, including actions taken to complete the repair, were recorded.

Upon LPG stove delivery, all intervention participants received the following behavioral support package: 1) a pledge by which they agreed to cook exclusively with the LPG stove during the trial, 2) training on how to safely operate the LPG equipment, and 3) behavioral messaging and materials to motivate exclusive LPG use and discourage biomass stove use. The messaging on exclusive LPG use was reinforced for any participants who showed evidence of traditional stove use during the trial. During reinforcement visits, participants were asked about their reasons for using the traditional stove and fieldworkers offered suggestions for alternatives.

### Stove Use Monitoring

We used a combination of stove use monitors (SUMs – data logging temperature sensors), surveys, and visual observations to assess stove use in our trial. Our SUMs monitoring efforts focused on ensuring that traditional stoves in intervention households were not used. This decision was driven by the evidence that traditional stove use must be nearly eliminated (<1 hour per week) to achieve clinically significant reductions in HAP.^15^ As such, we installed SUMs on all traditional stoves in intervention households. The SUMs recorded temperature readings every five minutes throughout the duration of the trial and were downloaded every two weeks. Temperature readings were used to flag cooking events based on temperature increases above identified thresholds that lasted at least five minutes (details on the algorithm are available in Quinn et al. 2021).^17^ In the event of device malfunction, we eliminated data that occurred after the error until the faulty SUM was repaired or replaced. We included only households that had at least two weeks of continuous SUMs data during pregnancy and/or the post-birth period (n=1,095 out of 1,584). We also installed SUMs in a subset of LPG stoves in intervention homes (n=276) and a subset of traditional stoves in control homes (n=214).

Surveys asking participants to report which stoves they had used in the prior 24 hours were administered with both intervention and control participants, twice during pregnancy (24-28 and 32-36 weeks of gestation) and three times post-birth (when the infant was approximately 3, 6, and 12 months old). Field staff conducted visual observations for evidence of traditional stove use at SUMs download visits in India, Peru, and Rwanda and at all visits in Guatemala. Any signs of recent traditional stove use (i.e., traditional stove being used during the visit, warm to the touch, or with fresh ashes) were recorded, in addition to whether the traditional stove in use had a SUMs installed in it.

### Data Analysis

Table 1 shows the indicators and metrics we used to evaluate fidelity and adherence to the intervention. We define fidelity (i.e., the extent to which the intervention was implemented as planned) as fulfillment of continuous LPG fuel delivery, timely repairs to LPG equipment, and behavioral reinforcement of exclusive LPG use. We define adherence as abandonment of traditional biomass stoves and adoption of the LPG stove among intervention participants, continued biomass stove use by control participants, and the extent to which these practices continued during temporary or permanent moves.

**Table 1.** Indicators and metrics used to measure intervention fidelity and adherence.

| Measure | Indicator | Metrics of assessment | Data source |
| --- | --- | --- | --- |
| Intervention Fidelity | Continuous LPG fuel delivery | # days between request for refill (or identification of refill need by fieldworkers) and LPG cylinder delivery | Surveys |
|  |  | % intervention participants reporting traditional stove use because they ran out of LPG |  |
|  | Timeliness of repairs | # and types of repairs made | Surveys |
|  |  | # days between problem identification and successful repair |  |
|  |  | # repairs per intervention participant |  |
|  | Behavioral reinforcement of exclusive LPG use | % intervention participants who reported using traditional stove due to problems or concerns with LPG | Surveys |
|  |  | # times intervention participants reported concerns with LPG |  |
|  |  | % intervention participants with observed traditional stove use who later received behavioral reinforcement |  |
|  |  | # days between observation of traditional stove use and reinforcement visit |  |
| Intervention Adherence | Disuse of traditional stoves in intervention households* | % of days with traditional stove use | SUMs |
|  |  | % intervention households with no traditional stove use |  |
|  |  | % intervention households with <1 day of traditional stove use per month |  |
|  |  | # days with traditional stove use per month |  |
|  |  | Average minutes of traditional stove use per day |  |
|  |  | % intervention households where traditional stove use was ever observed | Observations of traditional stove use |
|  |  | # observations of traditional stove use per intervention household |  |
|  |  | % observations indicating traditional stove use |  |
|  | LPG stove use by intervention participants | % intervention participants who reported exclusive LPG use across the study visits | Surveys |
|  |  | % days with LPG stove use | SUMs |
|  |  | # days with LPG stove use per month |  |
|  |  | Minutes of LPG stove use per day |  |
|  | Biomass stove use by control participants | % control participants who reported exclusive biomass use, mixed use of biomass and LPG, or exclusive LPG use across the study visits | Surveys |
|  |  | % days with biomass stove use | SUMs |
|  |  | Minutes of biomass stove use per day |  |
|  | Impact of temporary or permanent moves on adherence | % intervention participants who used biomass stoves in a new permanent home or temporary home | Surveys |
|  |  | % control participants who used exclusively clean fuel in new permanent or temporary home |  |
|  |  | # days in the new permanent or temporary home |  |

\* Note that we refer to intervention adherence at the household level given that the SUMs data do not distinguish which household member(s) are using the stove during flagged use events.

For each metric, we calculated the value during pregnancy, during the 12 months of follow-up after birth (infancy period), and across the full trial, by country and overall. We also calculated values for the follow-up that occurred before the onset of the global COVID-19 pandemic (‘pre-COVID,’ defined here as before March 17, 2020, which was the date across our study regions that best approximates the onset of major disruptions associated with local and national COVID-19 mitigation measures) and ‘post-COVID’ (March 17, 2020 or later) to assess any potential impact of COVID-19 mitigation strategies on our results. We additionally divided the post-COVID period into early (March 17-July 17, 2020) and late (after July 17, 2020) to assess for any differences between the initial months after the start of the COVID-19 pandemic (when nationally mandated lockdowns were instituted and new operating procedures were being developed) and later months after new routines were established. We used the Kolmogorov– Smirnov (K-S) test to compare differences by COVID-19 period and generalized estimating equations (GEE) in a logistic regression model with repeated measures, and accounting for overdispersion, to compare differences by birth period controlling for COVID-19 period.

Survey data were analyzed using StataSE version 15. SUMs data were analyzed using SAS version 9.4 and R version 4.2.2 (2022-10-31) – – “Innocent and Trusting”.

## RESULTS

### Study population

A total of 3,195 participants were enrolled and randomized between May 7, 2018 and February 29, 2020, 1,590 in the intervention group and 1,605 in the control group. Follow-up during pregnancy occurred between May 16, 2018 and September 17, 2020. Post-birth follow-up (the infancy period) occurred between September 21, 2018 and September 22, 2021. For the purposes of our analysis, we excluded six intervention participants who exited the study prior to receiving their LPG stove. Intervention participants were followed for nearly twice as many days in the infancy period compared to the pregnancy period (Table 2). Follow-up time was similar for control households (data not shown). Only 224 participants (14%) were still pregnant at the start of the global COVID-19 pandemic on March 17, 2020, with a remaining gestational period of a median 40.5 days (Q1, Q3: 14, 79) after March 17, 2020. A total of 1,324 participants (87%) contributed post-birth follow-up time for a median 237 days (Q1, Q3: 139.5, 345.5) after March 17, 2020.

**Table 2.** Average time in the study, stove repairs, and LPG deliveries for intervention participants during pregnancy (“preg”), the post-birth or infancy period (“infcy”), and total across the full trial (“full”).

|  | Guatemala |  |  | India |  |  | Peru |  |  | Rwanda |  |  | Total |  |  |
| --- | --- | --- | --- | --- | --- | --- | --- | --- | --- | --- | --- | --- | --- | --- | --- |
| Study period in relation to the baby’s birth | Preg | Infcy | Full | Preg | Infcy | Full | Preg | Infcy | Full | Preg | Infcy | Full | Preg | Infcy | Full |
| N Intervention Participants | 400 | 382 | 400 | 398 | 384 | 398 | 394 | 379 | 394 | 392 | 372 | 392 | 1584 | 1517 | 1584 |
| Median number of days in study period, (Q1, Q3) | 150<br>(130, 167) | 394<br>(383, 408) | 545<br>(521, 562) | 139<br>(120, 157) | 373<br>(366, 384) | 514<br>(494, 534) | 153<br>(133, 176) | 386<br>(377, 394) | 541<br>(517, 564) | 153<br>(135, 170) | 395<br>(377, 421) | 549<br>(522, 580) | 149<br>(129, 168) | 386<br>(374, 401) | 536<br>(511, 560) |
| # (%) of intervention participants who ever received a stove repair | 46<br>(12%) | 166<br>(43%) | 182<br>(46%) | 22<br>(6%) | 25<br>(7%) | 45<br>(11%) | 56<br>(14%) | 274<br>(72%) | 286<br>(73%) | 68<br>(17%) | 317<br>(85%) | 330<br>(84%) | 192<br>(12%) | 782<br>(52%) | 843<br>(53%) |
| Median (Q1, Q3) # of stove repairs received per participant, among those with any repair | 1.0<br>(1, 1) | 1.0<br>(1, 2) | 1.0<br>(1, 2) | 1.0<br>(1, 1) | 1.0<br>(1, 1) | 1.0<br>(1, 1) | 1.0<br>(1, 1) | 1.0<br>(1, 2) | 2.0<br>(1, 2) | 1.0<br>(1, 1) | 2.0<br>(1, 2) | 2.0<br>(1, 3) | 1.0<br>(1, 1) | 1.0<br>(1, 2) | 2.0<br>(1, 2) |
| Total repair visits made | 58 | 290 | 348 | 22 | 28 | 50 | 64 | 483 | 547 | 74 | 567 | 641 | 218 | 1368 | 1586 |
| Median (Q1, Q3) days between repair identification / request and successful repair | 0 (0, 0) | 0 (0, 0) | 0 (0, 0) | 0 (0, 0) | 0 (0, 0.5) | 0 (0, 0) | 0 (0, 0) | 0 (0, 0) | 0 (0, 0) | 0 (0, 0) | 0 (0, 0) | 0 (0, 0) | 0 (0,0) | 0 (0,0) | 0 (0,0) |
| Total LPG delivery visits made* | 4950 | 14673 | 19623 | 846 | 3423 | 4269 | 3293 | 9032 | 12325 | 1534 | 5101 | 6635 | 10623 | 32229 | 42852 |
| Median (Q1, Q3) days between delivery identification / request and successful delivery | 2 (1,3) | 2 (1,3) | 2 (1,3) | 5 (3,8) | 4 (3,6) | 4 (3,6) | 0 (0,0) | 0 (0,0) | 0 (0,0) | 1 (0,2) | 1 (0,3) | 1 (0,2) | 1 (0,2) | 1 (0,2) | 1 (0,2) |

\* Note that LPG cylinder sizes differed by country: 11.3 kg in Guatemala, 14.2 kg in India, 10 kg in Peru, and 12 or 15 kg in Rwanda.

### Intervention Fidelity

#### Continuous LPG fuel delivery

Across settings, a total of 42,852 visits were made to deliver LPG to households (10,623 during pregnancy and 32,229 post-birth) (Table 2). The median (Q1, Q3) time between request for or identification of the need for a refill and delivery of the LPG cylinder(s) was 1.0 days (0, 2). Median (Q1, Q3) time between refill request and delivery was highest in India (4 [3, 6] days), where local LPG distributors delivered LPG to households in coordination with the HAPIN team. Overall, 95.6% of LPG deliveries were completed within the 7-day target specified in the protocol. Time between refill request and delivery was lowest in Peru (median [Q1, Q3]: 0 [0, 0]), where fieldworkers often observed that cylinders were empty at SUMs download visits and installed a replacement cylinder at the same visit. Median (Q1, Q3) delivery times were similar during pregnancy (1.0 days; 0, 2) and post-birth (1.0 days; 0, 2) (Table 2), as well as pre-COVID (1.0 days; 0, 3) and post-COVID (1.0 days; 0, 2) (data not shown).

Overall, 26% of participants reported ever using their traditional stove because they ran out of LPG before a refill was delivered (Supplemental Table S1); however, despite this seemingly large percentage, the median (Q1, Q3) number of times these participants reported running out was low: 1 (1, 2). The percent of participants reporting running out of LPG was highest in Rwanda (67%), followed by Peru (27%) and Guatemala (11%). In India, no participants reported running out of LPG prior to a refill being delivered. Reports of running out of LPG were higher in the post-birth compared to the pre-birth period; however, this is likely due to the COVID-19 pandemic, which started when most participants were in the post-birth period. Figure 1 indicates that most reports of running out of LPG occurred in the first four months following the COVID-19 shutdowns in March 2020 (mostly from Peru and Rwanda). Instances of running out of LPG were less frequent in the later months of the COVID-19 pandemic once permissions were obtained and/or new operating procedures were established.

**Figure 1.**
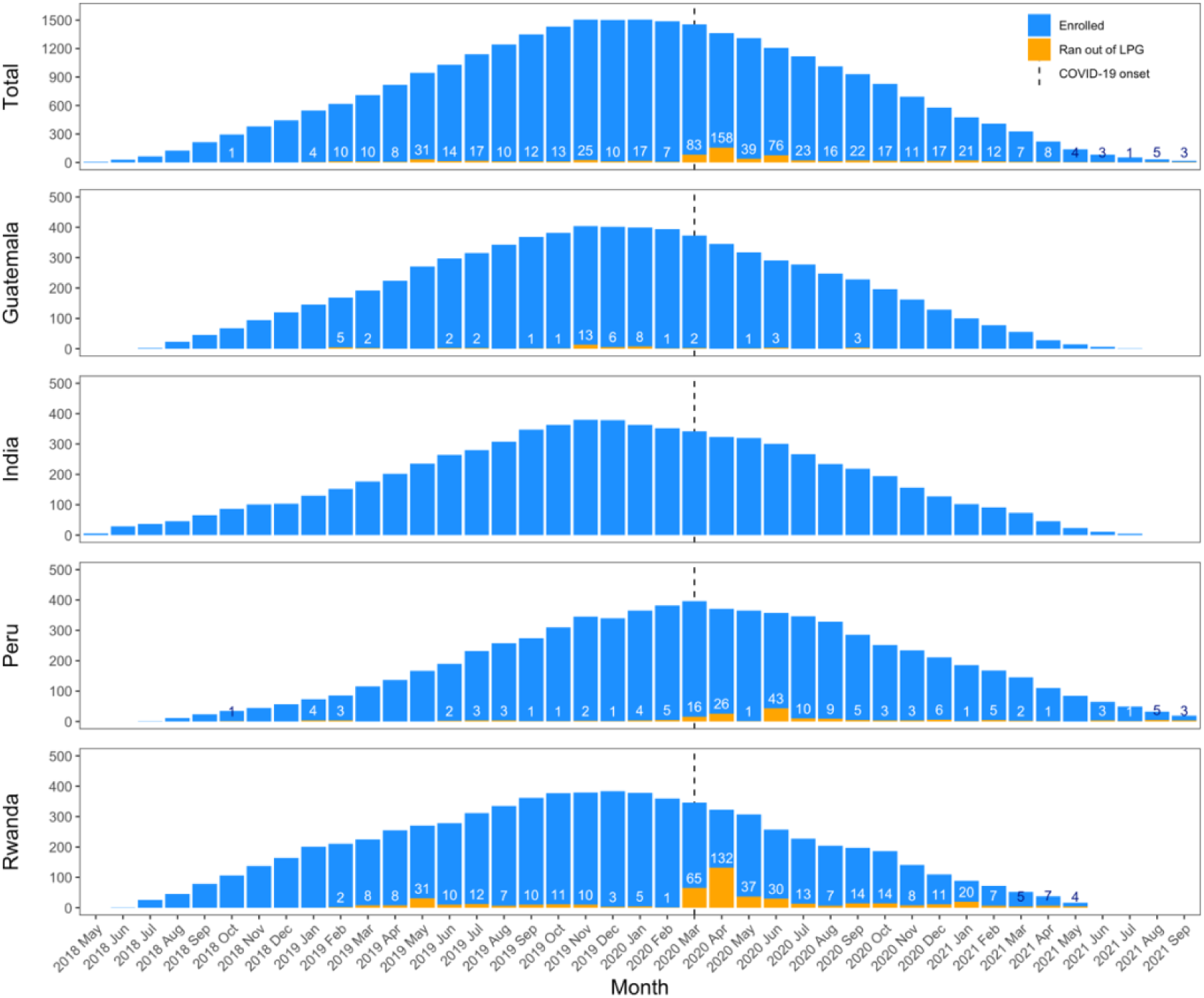
Number of participants reporting running out of LPG out of all enrolled intervention participants by month. Dashed line indicates onset of the global COVID-19 pandemic (March 17, 2020).

#### Timely repairs and assurance of LPG equipment functionality

Over the course of the study, a total of 1,586 repair visits were made to repair the LPG equipment, with most done the same day as the need for repair was identified (Table 2). Overall, 53% of participants required repairs, with a median (Q1, Q3) of 2.0 (1.0, 2.0) repairs per participant among those who ever received a repair (see Supplemental Table S2 for types of repairs). The need for repairs was lowest in India (11%), followed by Guatemala (46%), Peru (73%), and Rwanda (84%). There were no clear trends in the frequency of stove repairs over time (Supplemental Figure S1).

#### Behavioral reinforcement of exclusive LPG use

Overall, 43% of intervention participants reported problems, difficulties, or concerns with the LPG stove (including both equipment problems and user perceptions) at any behavioral reinforcement visit (Supplemental Table S1). Issues were more frequently reported in Rwanda (77% of participants ever reported a problem or difficulty), followed by Peru (54%), Guatemala (42%), and India (1%). The percent of participants reporting problems or difficulties was higher in the post-birth period compared to the pre-birth period in Guatemala (38% vs. 15%) and Rwanda (78% vs. 17%), but similar between periods in Peru (37% vs. 37%) and India (0% vs. 1%). Although nearly half of participants reported ever experiencing a problem, difficulty, or concern with the LPG stove, the median (Q1, Q3) number of reports per participant (among those with any reports) was low: 2 (1, 3).

Reported reasons for occasional use of the traditional stove were similar in pre-birth and post-birth periods (data not shown), except for running out of LPG, which was more common in the post-birth compared to the pre-birth period in Guatemala, Peru, and Rwanda (likely driven by the higher frequency of running out of LPG in the early months of the COVID-19 pandemic, see Figure 1). In Rwanda, reported challenges with cleaning or maintaining the LPG stove or the stove not functioning correctly also increased post-birth compared to pre-birth. Few problems with the LPG stove were reported in India. The most commonly reported reasons for traditional stove use by country across the full trial are shown in Figure 2.

**Figure 2.**
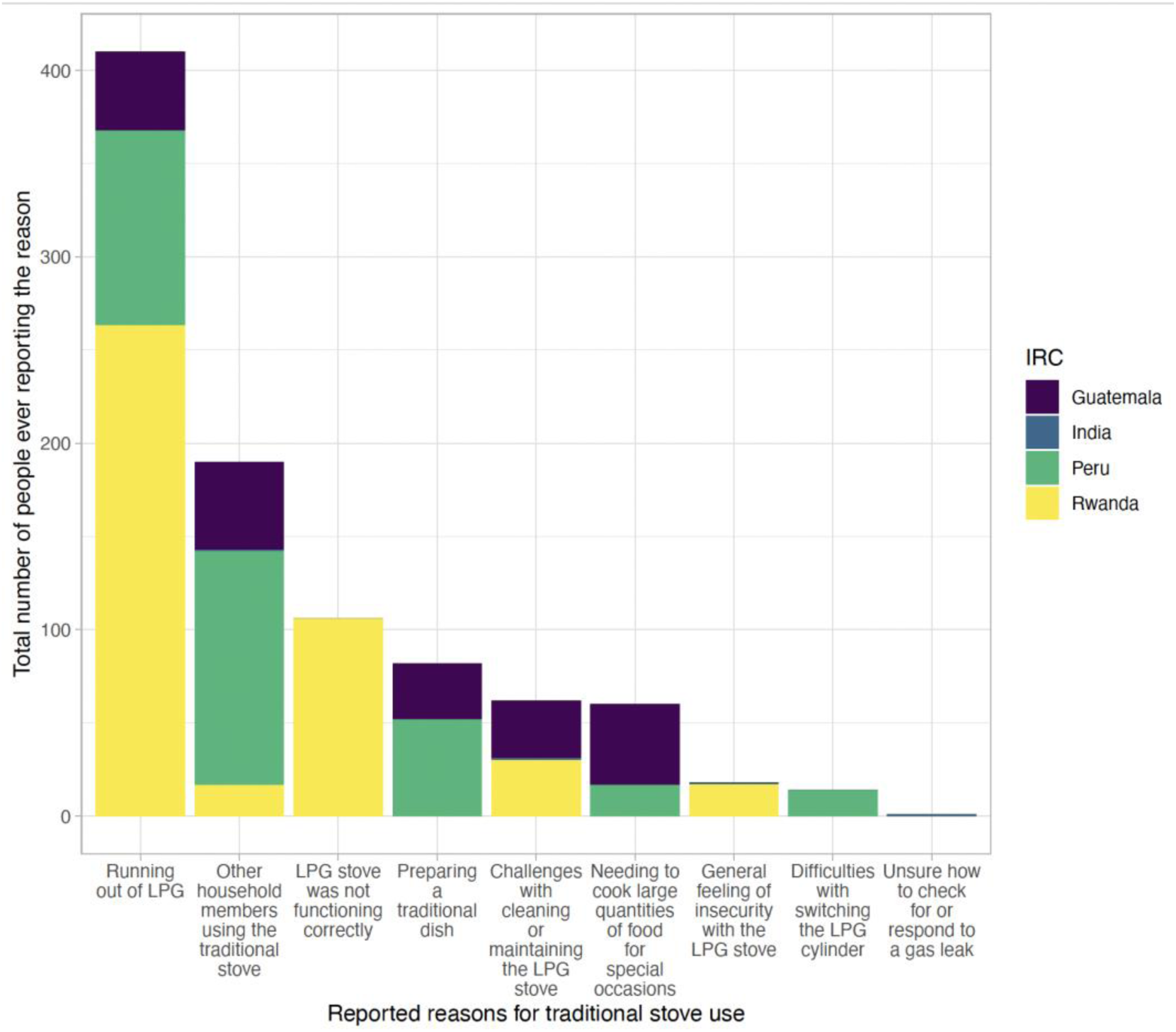
Top reasons for traditional stove use (including the top five reported reasons from each country), showing the total number of people who ever reported each reason by country. Bars for India are very small given that very few people ever reported a problem or concern about LPG (n=3).

Eighty-nine percent (n= 1,346) of observations of traditional stove use were followed up with a behavioral reinforcement visit (Supplemental Table S3). The median (Q1, Q3) number of days between observation of traditional stove use and the reinforcement visit was 9 (0, 56).

### Intervention Adherence

#### Disuse of traditional stoves in intervention households

Of the 1,087 intervention participants who retained a traditional stove in their household during pregnancy following LPG stove installation, 96% (n=1,039) had their traditional stove(s) successfully monitored by a SUM (Supplemental Table S4). Similarly, of the 1,020 intervention participants who had a traditional stove in their household after birth (including some households who re-installed a traditional stove that had been destroyed during pregnancy), 86% (n=877) had their traditional stove(s) successfully monitored by a SUM (see Supplemental Table S4 for potential reasons for missing SUMs data). A total of 218 participants had SUMs data only in pregnancy, 56 only post-birth, and 821 in both pregnancy and post-birth, for a total of 1,095 intervention households with traditional stoves monitored by SUMs at some point during the trial. In households with SUMs installed, the SUMs successfully recorded data for the majority of follow-up time: 84.3% of days between each participant’s date of LPG stove delivery and their exit date were monitored by SUMs (Table 3). The proportion of follow-up time monitored by SUMs was lower in Guatemala because many participants destroyed their traditional stove and thus only had SUMs data from a small percentage of their total time in the study.

**Table 3.**
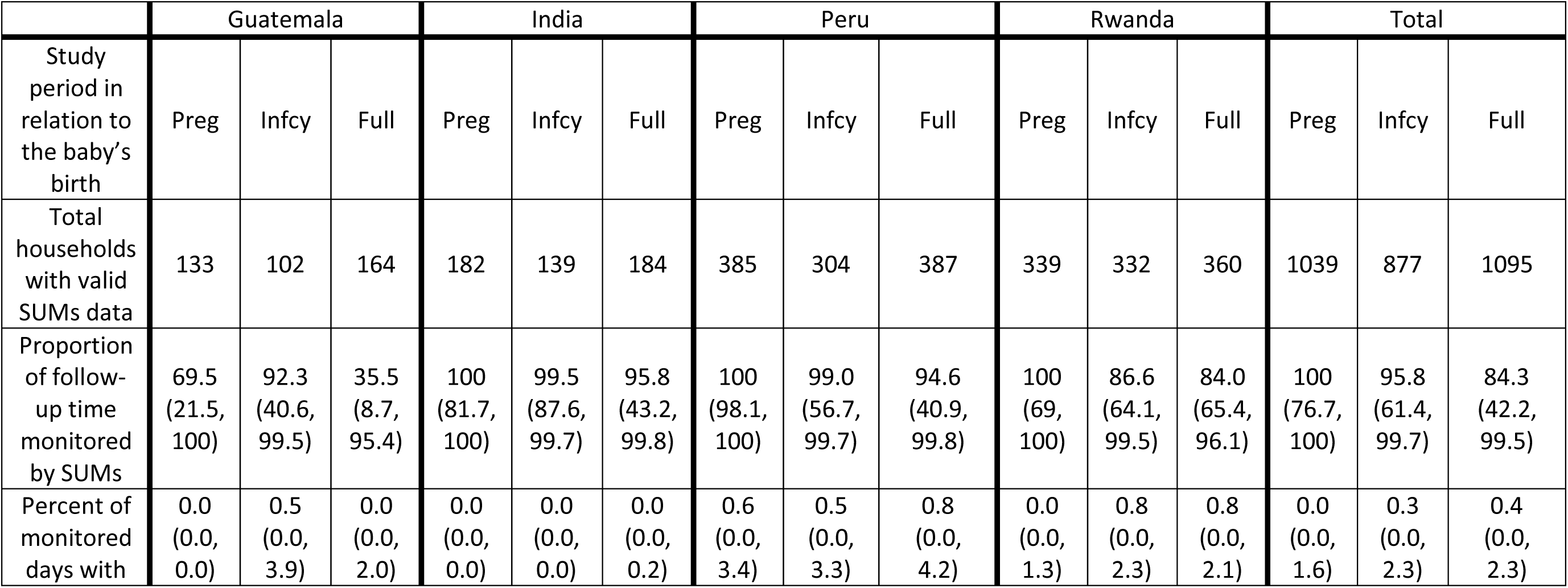
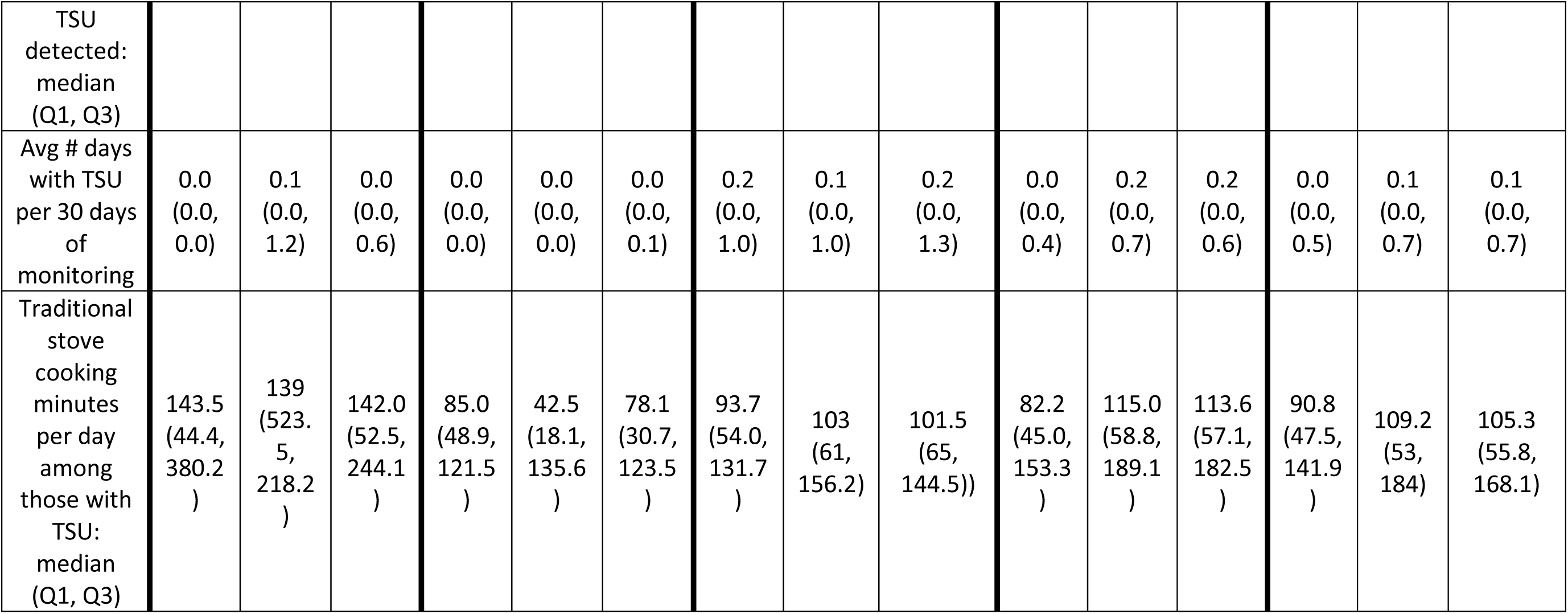
Traditional stove use (TSU) by intervention participants based on SUMs data during pregnancy (“preg”), the post-birth or infancy period (“infcy”), and total across the full trial (“full”).

Over the full trial, intervention households with SUMs installed rarely used their traditional stove, for a median (Q1, Q3) of 0.4% (0.0%, 2.3%) of monitored days (Table 3, Supplemental Table S5). The median percent of days in which the traditional stove was used was slightly higher in Peru (0.8%) and Rwanda (0.8%) than in Guatemala (0.0%) and India (0.0%), but consistently low overall. Although a few intervention households used their traditional stoves more frequently (Figure 3; Supplemental Figures S2, S3), 85% either removed their traditional stove (29%), never used their traditional stove (29%), or used their traditional stove on less than one day out of every 30 days (27%) (Figure 4). Households with SUMs had a median (Q1, Q3) of 0.1 (0.0, 0.7) days with TSU per 30 days of monitoring (Table 3). On days with any traditional stove use, intervention participants spent a median (Q1, Q3) of 105.3 (55.8, 168.1) minutes cooking with the traditional stove. Results were similar in the subset of households with a non-pregnant adult woman enrolled (aged 40-79 years; n=143 with SUMs monitoring): traditional stove use was detected on a median (Q1, Q3) of 0.4% (0.0%, 3.6%) of monitored days and 0.1 days (0.0, 1.1) per 30 days of monitoring (Supplemental Table S6).

**Figure 3.**
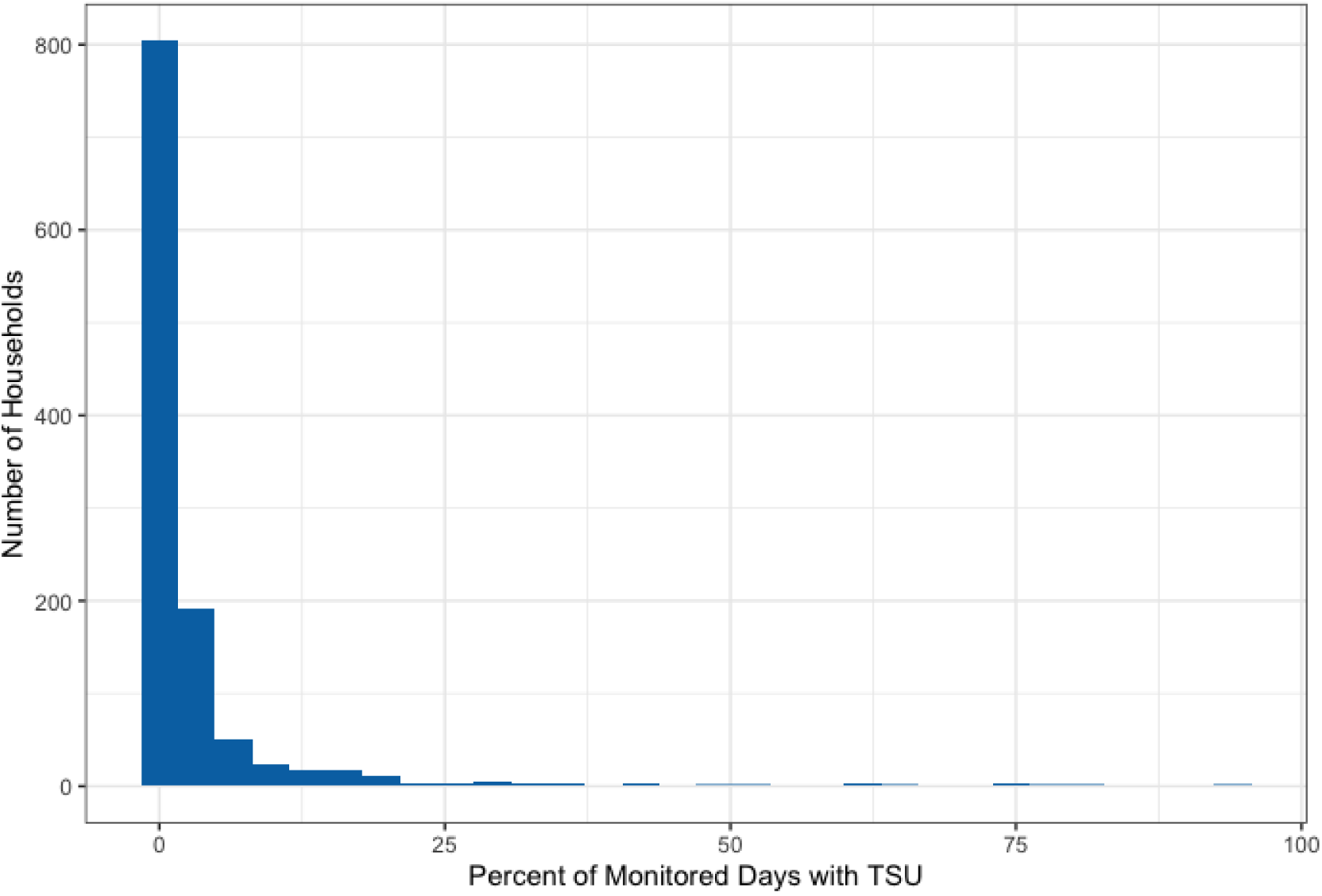
Frequency of the percent of stove-use-monitored days in which traditional stove use (TSU) was detected via stove use monitors (SUMs) in intervention households over the full trial period.

**Figure 4.**
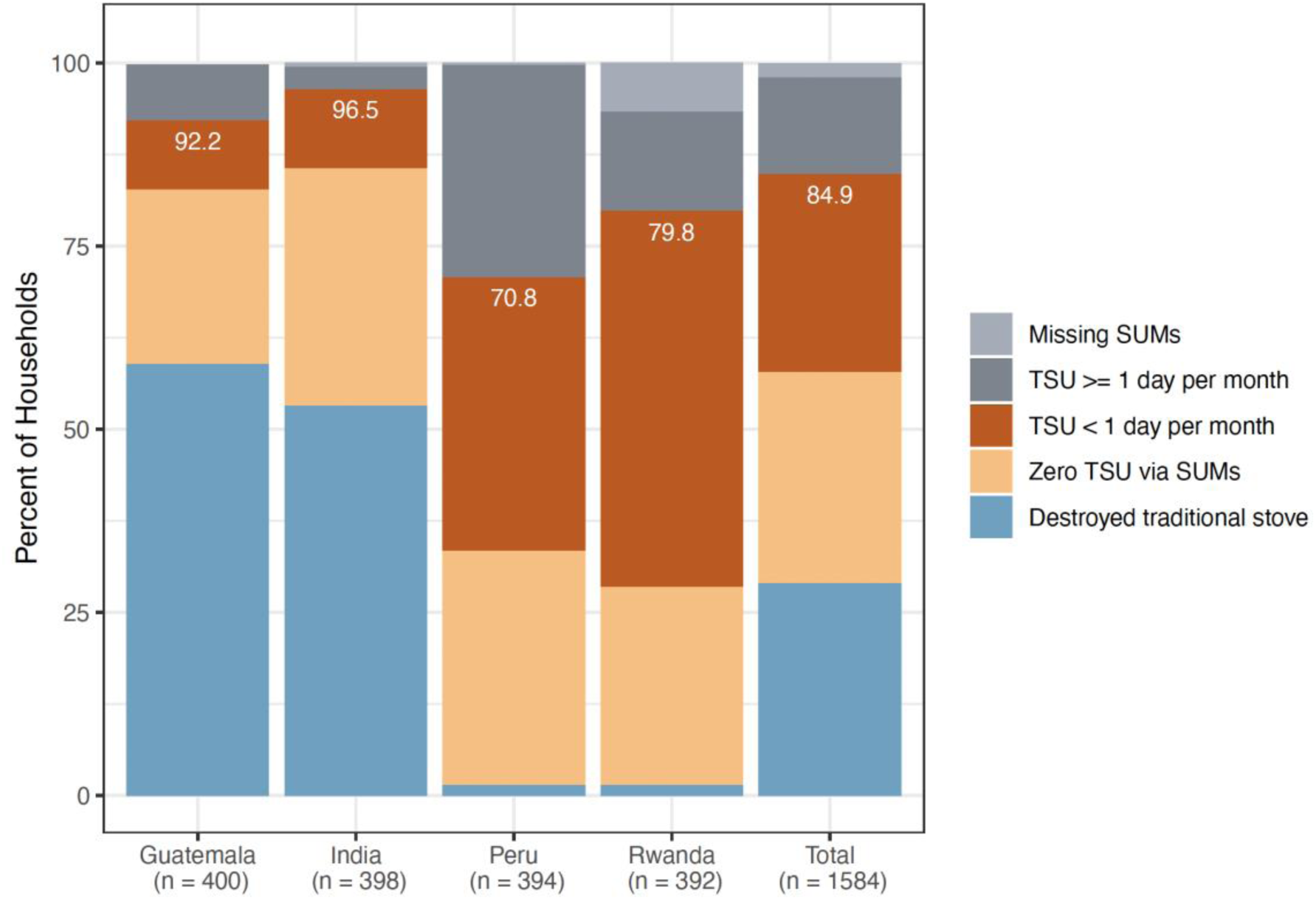
Percent of intervention households who destroyed their traditional stove at LPG stove installation and have no subsequent SUMs data (blue), had zero traditional stove use (TSU) according to SUMs data (sepia), or had less than one day of TSU per month (red), by country and overall. Percentages displayed in white represent the total percent of participants falling into either of these three categories. Grey bars represent participants with one or more day with traditional stove use per month (dark grey) or who retained their traditional stove but are missing SUMs data (light grey).

After controlling for COVID-19, there were no statistically significant differences in traditional stove use during the pregnancy period (median 0.0% [Q1, Q3: 0.0%, 1.6%]) as compared to the infancy period (median 0.3% [Q1, Q3: 0.0%, 2.3%]) (odds ratio=1.11 [CI: 0.95-1.30], *p*=0.1866) (Table 3, Figure 5). The onset of the COVID-19 pandemic was associated with a statistically significant increase in traditional stove use among some users, but medians remained the same at 0.0% during the pre-COVID-19 period (Q1, Q3: 0.0%, 1.6%) and 0.0% during the post-COVID-19 period (Q1, Q3: 0.0%, 3.4%) (Supplemental Table S7) (KS test D=0.12, p<0.001). More people had a significantly greater proportion of days with traditional stove use in the first four months after the start of the COVID-19 pandemic (median: 0.0%; Q1, Q3: 0.0%, 3.5%) compared to later months of the COVID-19 period (median: 0.0%; Q1, Q3: 0.0%, 1.5%) (Figure 5, Supplemental Table S8) (KS test D=0.12, p=0.001).

**Figure 5.**
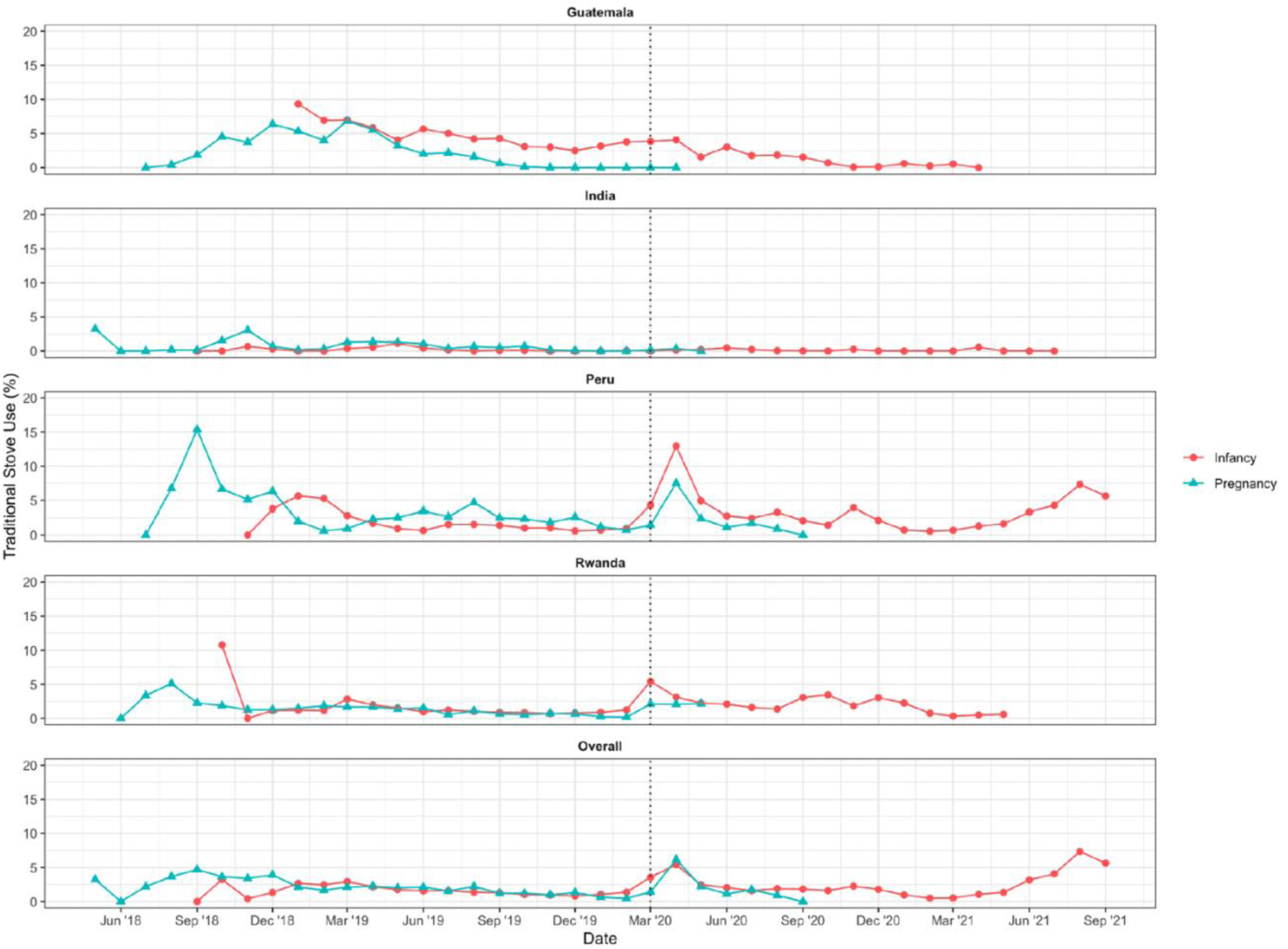
Percent of monitored days with traditional stove use as recorded by SUMs by calendar month during pregnancy and post-birth (“infancy”) periods, by country and overall among intervention participants. The dashed line indicates the onset of the COVID-19 pandemic, which began March 17, 2020 (shown at the March ‘20 position given that all March data was aggregated into one point).

Nearly all intervention participants (1,575/1,584 or 99%) received at least one visit in which fieldworkers looked for signs of any traditional stove use, with a median (Q1, Q3) of 29 (23, 34) observation visits per participant over the course of the trial (Supplemental Table S9). A total of 55,241 observation visits were made, of which only 1,506 (3%) indicated recent traditional stove use (Supplemental Table S3). Observations indicating traditional stove use were rare in India (only 5 participants [1.3%] were ever observed to have used their traditional stove recently) (Supplemental Table S9). Although more participants were observed to have ever used their traditional stove in Guatemala (n=224; 56%), Peru (n=202; 51%), and Rwanda (n=229; 58%), the median (Q1, Q3) number of traditional stove use observations per participant among those with any observed use was low: 2.0 (1, 3) observations per participant (excluding India). Overall, among participants with any observations of traditional stove use, traditional stove use was only observed in a median (Q1, Q3) of 4.5% (3%, 9%) of observation visits.

Most traditional stoves that were observed to have been recently used had a SUMs installed (64%) (Supplemental Table S3), suggesting that most traditional stove use events were captured and reflected in the SUMs data. For the 547 households with no SUMs data during pregnancy and the 640 households with no SUMs data post-birth (i.e., due to removal of the traditional stove or missing SUMs data), observations for traditional stove use were completed in 531 (97%) and 639 (99%) of households during the pregnancy and post-birth periods, respectively (Supplemental Table S10). Out of a median (Q1, Q3) of 10 (6, 16) observations completed in those households during pregnancy and 22 (18, 47) post-birth, the median (Q1, Q3) number of traditional stove uses observed was 0 (0, 0) in both periods. This indicates that traditional stove use was also low or non-existent in households lacking SUMs data.

#### Use of the LPG stove in intervention households

In the subset of 276 intervention households who had SUMs installed on their LPG stoves, LPG use was detected on a median (Q1, Q3) of 95.2% (87.8%, 98.6%) of all monitored days. The LPG stove was used for a median (Q1, Q3) of 28.6 (26.3, 29.6) days per 30 days of monitoring and 260.9 (204.2, 325.4) minutes of use per day. Self-reported data showed that in the majority of pregnancy and post-birth survey visits (96.6%), intervention participants reported using LPG exclusively in the preceding day (Figure 6). Nearly 90% of participants reported using LPG exclusively in all survey visits, and 99.3% of participants reported exclusive LPG use in 75% or more of survey visits.

**Figure 6.**
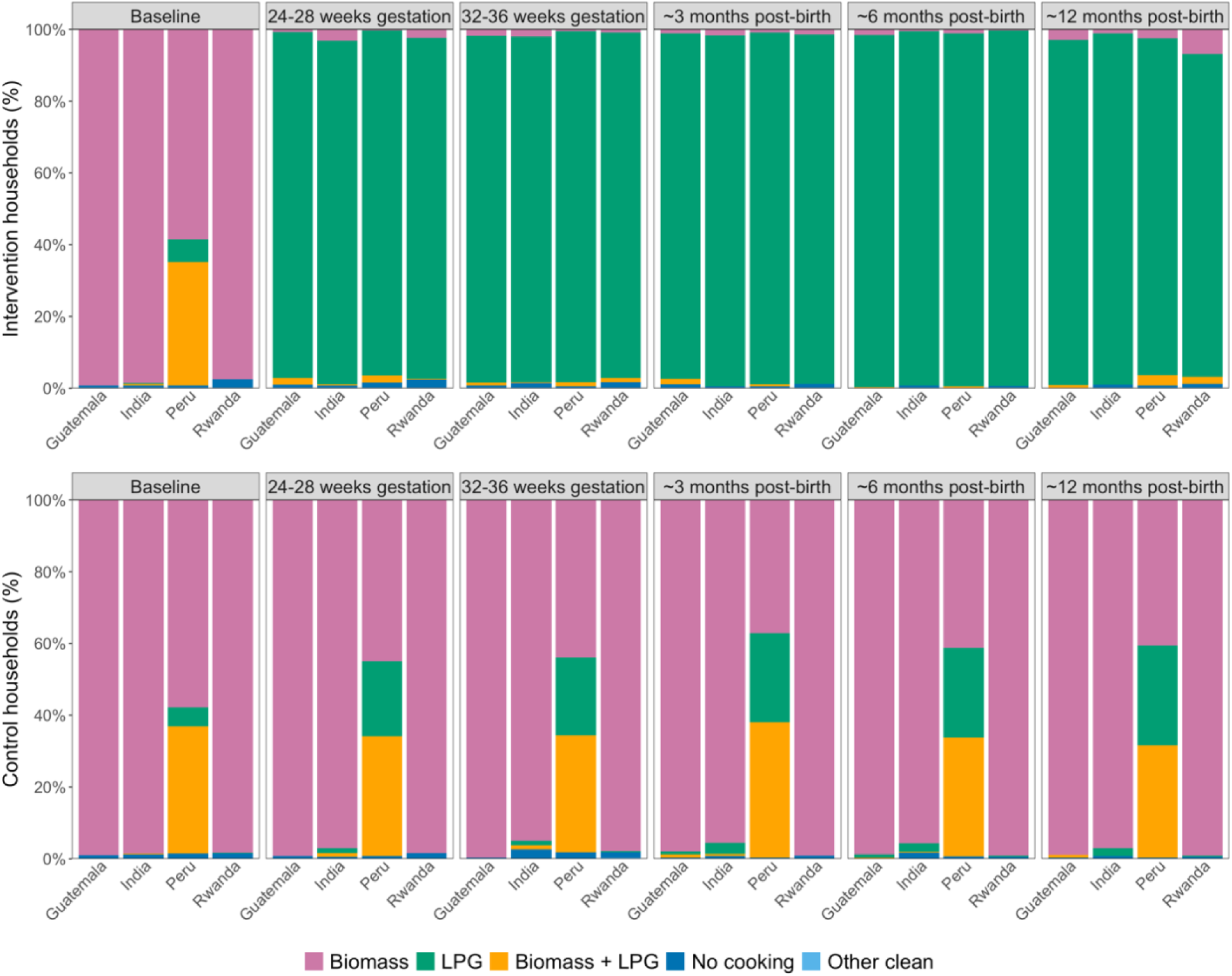
Self-reported stove use in the 24-hours preceding the survey at baseline, 24-28 and 32-36 weeks gestation, and 3, 6, and 12 months post-birth among intervention and control participants. Pink represents exclusive biomass use; green, exclusive LPG use; orange, mixed use of biomass and LPG; dark blue, no cooking; and light blue, cooking with other clean technologies such as electric stoves.

#### Use of biomass and LPG stoves in control households

Across all study visits in pregnancy and post-birth, control participants in Guatemala, India, and Rwanda reported exclusive use of biomass (97.9%) or a mix of LPG and biomass (0.3%) (Figure 6). In Peru, control participants reported some use of biomass in 76% of all visits (42% exclusive and 34% mixed); although 24% of all visits indicated exclusive LPG use, only 17 participants (4.5%) reported exclusive LPG use at every survey visit. Across all countries, approximately 84% of control participants reported using biomass in all survey visits, and 91.1% reported using biomass in 75% or more of survey visits. In the subset of control households with SUMs installed in traditional stoves (n=214), traditional stove use was recorded on a median (Q1, Q3) of 69.3% (43.4%, 87.8%) of monitored days and used for 245.7 (165.5, 334.4) minutes per day of use.

#### Impact of temporary or permanent moves on adherence

Overall, 22% of intervention participants and 23% of control participants moved to either a temporary or permanent home during the trial (Supplemental Table S11). Most intervention participants continued using LPG exclusively in their new or temporary home (95%), and most control participants continued using biomass either primarily or secondarily in their new or temporary home (96.5%).

## DISCUSSION

The research questions that the HAPIN trial sought to answer required an understanding of the intervention fidelity and adherence. To achieve the highest possible adherence, special steps were undertaken, including free LPG stoves and fuel delivered directly to the home, timely repairs, locally tailored behavioral messaging, and comprehensive stove use monitoring. The trial achieved high fidelity to implementation of these intervention components, which were successful in contributing to high intervention adherence by intervention participants throughout the 18-month trial period, while most control participants continued to use primarily biomass fuels for cooking. The level of adherence to LPG stove use achieved among the intervention participants in the HAPIN trial aligns with that observed in a smaller scale randomized controlled LPG intervention in Peru^13^, and is much higher than has been reported in other clean cooking trials^8, 18, 19^.

Our study did not find that adherence to exclusive LPG use was significantly different during pregnancy compared to post-birth. This contrasts with Carrión et al. (2020), who found that LPG stove use was higher during pregnancy compared to during the infant’s first year of life in a randomized controlled trial in Ghana (95.4% reported using the LPG stove for the main meal the previous day during pregnancy, compared to 82.8% over one year post-birth).^18^ Although we found that traditional stove use was slightly higher post-birth compared to during pregnancy, this was likely an effect of COVID-19 given that most participants were already in or near the post-birth follow-up period when the COVID-19 shutdown began. Indeed, after controlling for the COVID-19 pandemic, differences in adherence between pregnancy and post-birth periods were non-significant.

Notably, we found that participants in India had higher adherence (both in terms of traditional stove use flagged by SUMs and by observations), fewer reports of problems or concerns with LPG, and fewer documented stove repairs than in Peru, Guatemala, or Rwanda. Greater exclusivity of LPG use in India may have been driven by enhanced awareness of and motivation to use LPG in the country due to national policies, such as the Pradhan Mantri Ujjwala Yojana (PMUY) LPG subsidization program for households below the poverty line.^20, 21^ Additionally, because LPG was delivered by local distributors in India, they may have made simple stove or tank repairs or addressed small concerns that were not reported to the study team, thus resulting in a lower number of documented repairs and reported problems in India compared to other countries where study staff made LPG deliveries.

Our results indicate that the COVID-19 pandemic temporarily impacted the timely delivery of LPG (mainly in Peru and Rwanda), contributing to slightly higher traditional stove use during this period. Nonetheless, traditional stove use remained extremely low overall. Our findings suggest that the impacts of the COVID-19 pandemic were mainly limited to the first four months of the pandemic, and that this did not jeopardize the high rate of fidelity and adherence achieved.

Our field teams’ frequent observations for traditional stove use aligned with the adherence data obtained from the SUMs. Across over 55,000 observations made, only 3% indicated any traditional stove use and only 1% indicated use of a traditional stove that did not have a SUMs installed. Additionally, in households lacking SUMs data either due to removal of the traditional stove or missing data, observations rarely indicated any traditional stove use. Also, our tracking of household moves showed that most participants continued to use the stove indicated by their assigned randomization group.

Factors driving the high rates of adherence to LPG use observed among our intervention participants may include the study’s rapid delivery of free LPG in response to refill needs (with deliveries completed in a median of 1 day from the time of request), provision of behavioral reinforcement to participants who used their traditional stove (89% of traditional stove use events were followed up with behavioral reinforcement), and timely completion of repairs to LPG equipment (with repairs completed in a median of 0 days from the time that the need for a repair was identified). Additionally, the extensive training and education provided to participants at the beginning of the study and the participants’ completion of pledges to use the LPG stove for all household cooking needs may have also motivated high adherence, as reported in Quinn et al. (2021).^17^ Another research trial in Peru that also reported high adoption of LPG similarly found that behavioral training and reinforcement, as well as support from fieldworkers, motivated LPG use.^22^

Overall, we found that occasional traditional stove use was mainly driven by factors outside of the participants’ control (i.e. running out of LPG, other household members using the traditional stove, or problems with the LPG stove); cultural traditions and taste preferences were less commonly cited as reasons for using the traditional stove, suggesting that such factors, which have been previously believed to influence adoption decisions^23^, are no longer significant when financial and structural barriers to LPG use are removed. Our study found that other household members using the traditional stove was among the top five reasons for traditional stove use across all four countries. This indicates that a large portion of the traditional stove use flagged in our study may have been done by someone else in the household other than the pregnant woman/mother. Thus, our adherence estimates may overestimate the extent to which the pregnant women/mothers were using and directly exposed to the emissions of the traditional stoves. Additionally, as similarly reported by Williams et al. (2020)^22^, LPG delivery delays, resulting in participants running out of LPG, also triggered some traditional stove use. Problems with the LPG stove also resulted in some traditional stove use while waiting for repairs or behavioral support on specific issues.

We found a fairly consistent need for stove and equipment repairs over time, despite the fact that stove models selected were all expected to be of high quality, durable, and compatible with local cooking needs based on formative research and pilot results.^16, 24^ This indicates that programs seeking to distribute or promote LPG stoves must ensure the infrastructure is available and affordable for continued repair and maintenance to ensure continued stove functionality, as highlighted by Gould et al. (2018).^25^ However, the need for complete stove replacement was rare, suggesting that with continuous maintenance and small repairs, the stoves were sufficiently durable to last throughout the trial.

Strengths of our study include use of multiple sources of adherence data to assess consistency (i.e., direct-reading instruments, self-report, and observations), bi-weekly visits to households to verify placement and download SUMs data to limit any data loss, checking LPG stoves twice monthly to ensure functionality, and systematic tracking of intervention implementation (i.e., LPG delivery, repairs, and behavioral reinforcement). However, several limitations to our fidelity and adherence data should be noted. First, although we deployed high quality SUMs with continuous tracking through an online dashboard^26^, it is possible that some traditional stove use events may have been missed or that some non-events may have been erroneously flagged as use. Additionally, some participants may have used unmonitored traditional stoves not observed by fieldworkers. Observations and stove use surveys typically occurred on weekdays, meaning weekend use may have been less likely to be captured through these methods (although SUMs were continuously in place and thus captured both weekday and weekend use). Our extensive follow-up and examination of multiple adherence data sources suggests that unmonitored traditional stove use was infrequent. Second, national shutdowns due to the COVID-19 pandemic, which began partway through our trial, necessitated some adjustments and adaptations to trial implementation. Although we adapted study procedures and data collection methods to align with global safety protocols as described in Simkovich et al. (2021)^27^, there were some delays and interruptions to fidelity as new procedures were established. Despite these, however, overall fidelity and adherence remained high.

## CONCLUSION

High fidelity to timely delivery of LPG, repairs, and behavioral reinforcement contributed to near-exclusive LPG use among intervention participants across four distinct HAPIN study sites. Results provide insight into the level of LPG adoption that can be achieved with intensive economic, behavioral, fuel delivery, and maintenance support. This level of fidelity and adherence helps us to answer the health outcome research questions posed by the HAPIN trial. However, the same should not be assumed from clean fuel interventions delivered programmatically that rely on household members to bear most or all of the costs of the stoves and fuel, where access is challenging, behavior change messaging limited, and stove use monitoring absent.

### Institutional Review Board Statement

The study protocol was reviewed and approved by institutional review boards (IRBs) or Ethics Committees at Emory University (00089799), Johns Hopkins University (00007403), Sri Ramachandra Institute of Higher Education and Research (IEC-N1/16/JUL/54/49) and the Indian Council of Medical Research – Health Ministry Screening Committee (5/8/4-30/(Env)/Indo-US/2016-NCD-I), Universidad del Valle de Guatemala (146-08-2016) and Guatemalan Ministry of Health National Ethics Committee (11-2016), Asociación Beneficia PRISMA (CE2981.17), the London School of Hygiene and Tropical Medicine (11664-5) and the Rwandan National Ethics Committee (No.357/RNEC/2018), and Washington University in St. Louis (201611159). The study was registered with ClinicalTrials.gov (Identifier NCT02944682). Informed consent was obtained from all participants.

## Data Availability

The data that support the findings of this study are available from the corresponding author upon reasonable request.

## Acronyms and abbreviations

Intvn: Intervention group
Cntrl: Control group
HAP: Household Air Pollution
HAPIN: Household Air Pollution Intervention Network trial
LPG: Liquefied petroleum gas
K-S: Kolmogorov–Smirnov test
Preg: Follow-up that occurred during the pregnancy period, from delivery of the LPG stove through birth of the child
Infcy: Follow-up that occurred during the post-birth or “infancy” period, from birth through the child’s first birthday
Pre-COVID-19: Period of follow-up occurring prior to the onset of the global COVID-19 pandemic, between May 7, 2018 and March 16, 2020
Post-COVID-19: Period of follow-up occurring after the onset of the global COVID-19 pandemic, between March 17, 2020 and the end of the trial (September 22, 2021)
PM_2.5_: Fine particulate matter
SUMs: Stove use monitors
TS: Traditional stove
TSU: Traditional stove use
WHO: World Health Organization

## Acknowledgements

The HAPIN trial was funded by the U.S. National Institutes of Health (NIH; cooperative agreement 1UM1HL134590) in collaboration with the Bill & Melinda Gates Foundation (OPP1131279). A multidisciplinary, independent Data and Safety Monitoring Board (DSMB) appointed by the National Heart, Lung, and Blood Institute (NHLBI) monitored the quality of the data and protected the safety of patients enrolled in the HAPIN trial. NHLBI DSMB: Catherine Karr (Chair), Nancy R. Cook, Stephen Hecht, Joseph Millum, Nalini Sathiakumar, Paul K. Whelton, Gail Weinmann, and Thomas Croxton (Executive Secretaries). Program Coordination: Gail Rodgers, Bill & Melinda Gates Foundation; Claudia L. Thompson, National Institute of Environmental Health Science; Mark J. Parascandola, National Cancer Institute; Marion Koso-Thomas, Eunice Kennedy Shriver National Institute of Child Health and Human Development; Joshua P. Rosenthal, Fogarty International Center; Concepcion R. Nierras, NIH Office of Strategic Coordination Common Fund; Katherine Kavounis, Dong-Yun Kim, Antonello Punturieri, and Barry S. Schmetter, NHLBI. The investigators would like to thank the Drs. Patrick Breysse, Donna Spiegelman, and Joel Kaufman (members of the advisory committee) for their valuable insight and guidance throughout the implementation of the trial. We also wish to acknowledge all the research staff and study participants for their dedication to and participation in this important trial. The findings and conclusions in this report are those of the authors and do not necessarily represent the official position of the US National Institutes of Health or Department of Health and Human Services. Under the grant conditions of the Bill & Melinda Gates Foundation, a Creative Commons Attribution 4.0 Generic License has already been assigned to the Author Accepted Manuscript version that might arise from this submission. The data that support the findings of this study are available from the corresponding author upon reasonable request.

## Supplemental Materials

**Figure S1.**
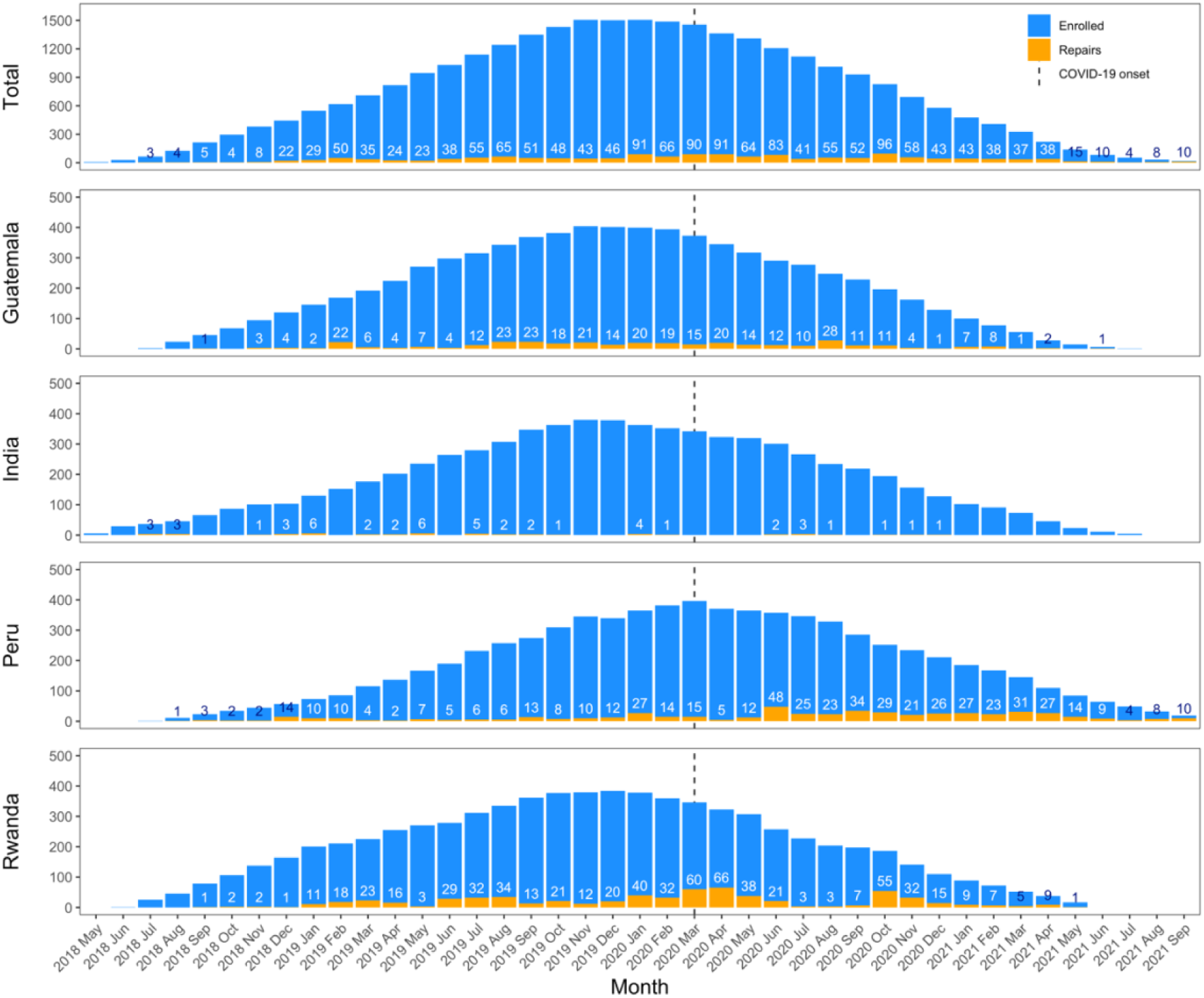
Number of participants receiving a stove repair out of all enrolled participants by month. Dashed line indicates onset of the global COVID-19 pandemic (March 17, 2020).

**Figure S2.**
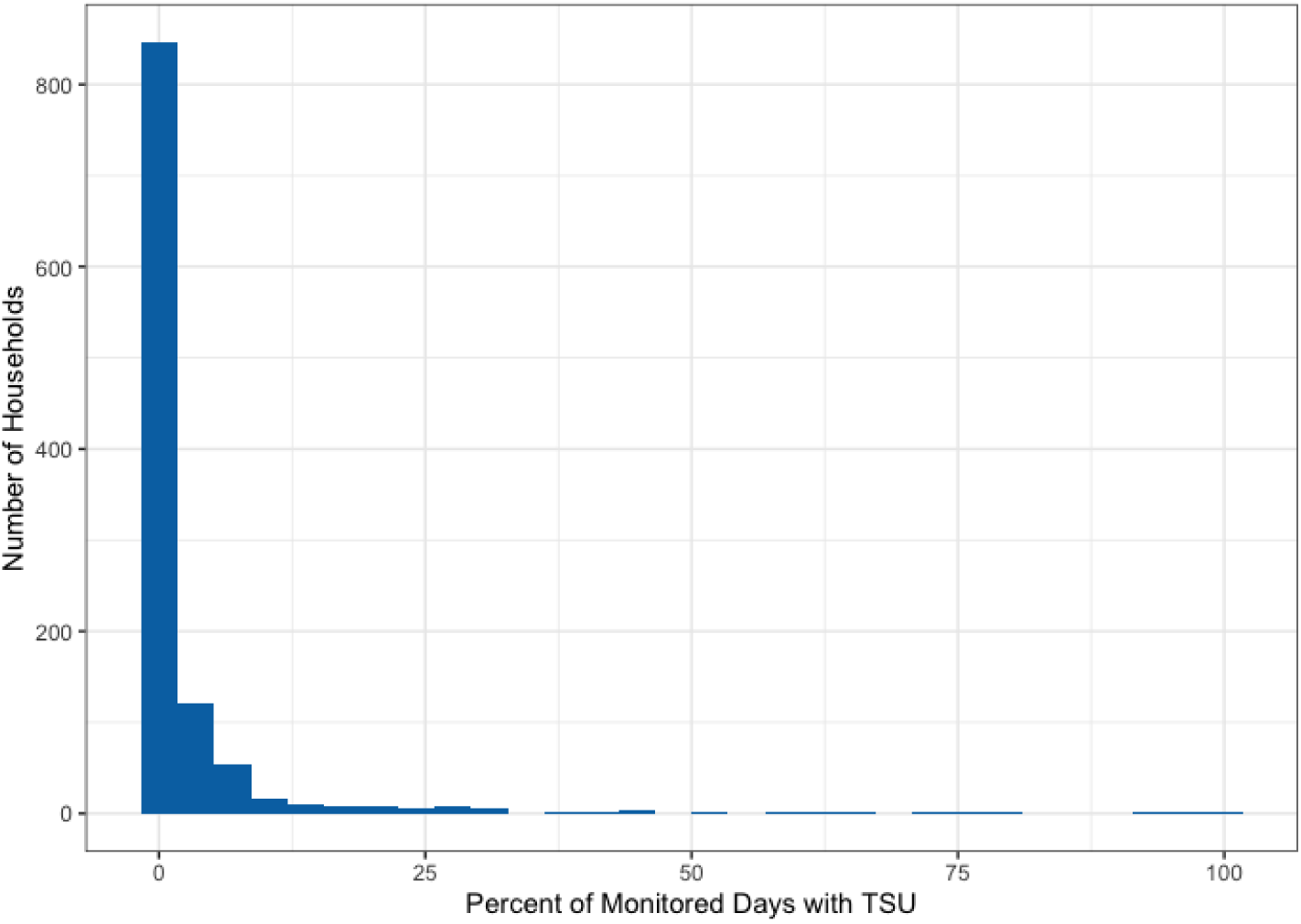
Frequency of the percent of stove-use-monitored days in which traditional stove use (TSU) was detected via stove use monitors (SUMs) in intervention households during the pregnancy period.

**Figure S3.**
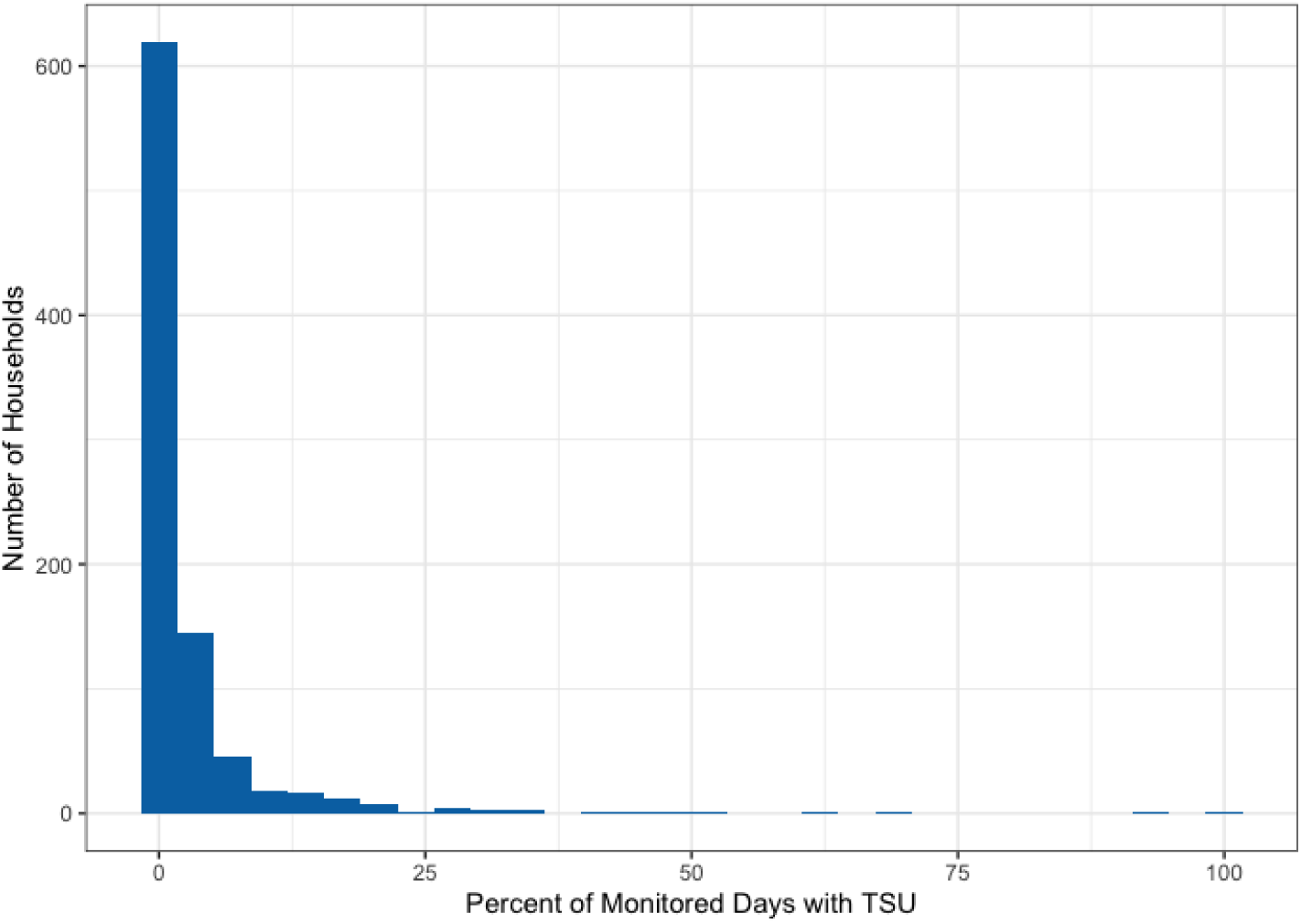
Frequency of the percent of stove-use-monitored days in which traditional stove use (TSU) was detected via stove use monitors (SUMs) in intervention households during the post-birth or infancy period.

**Table S1.**
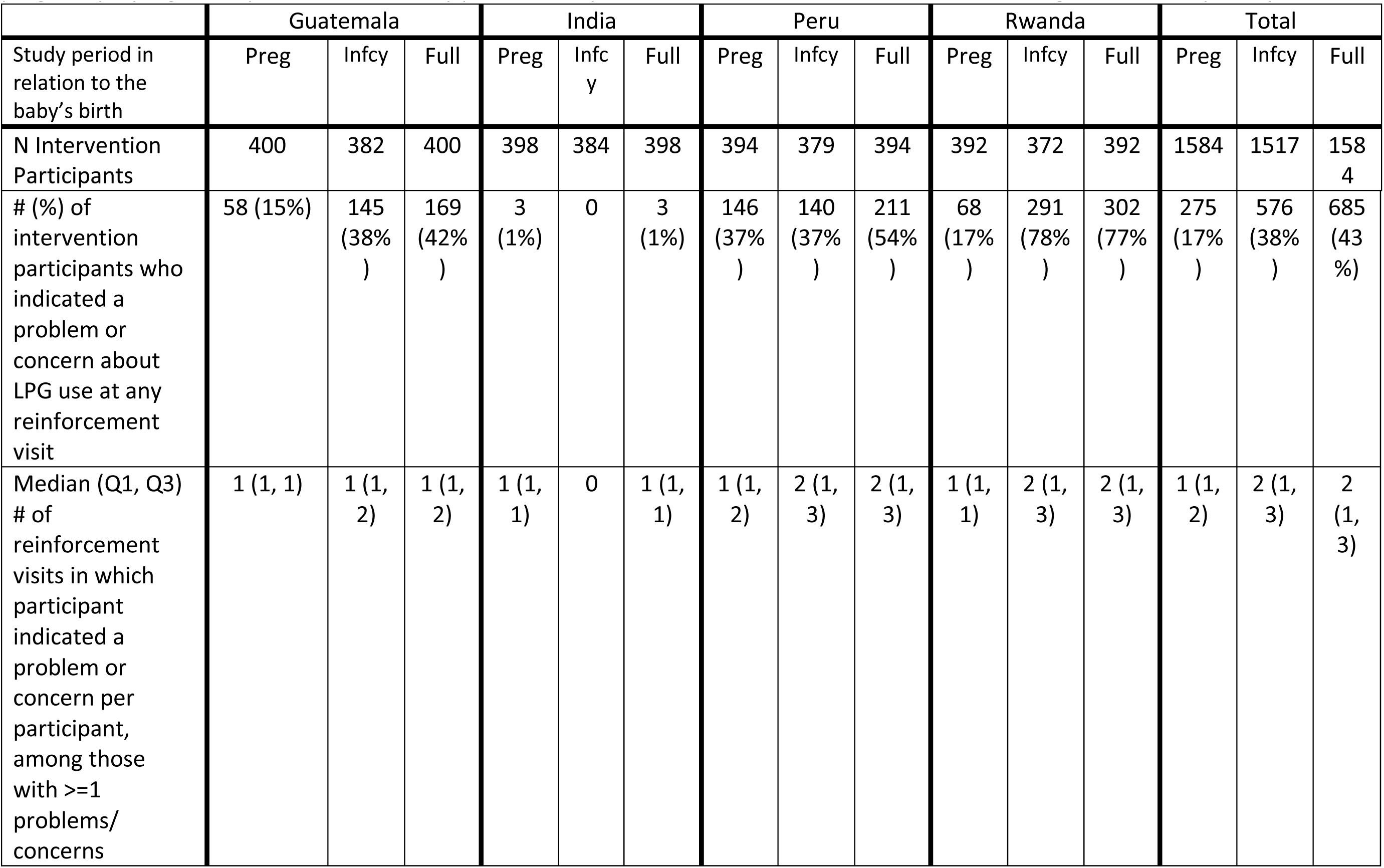
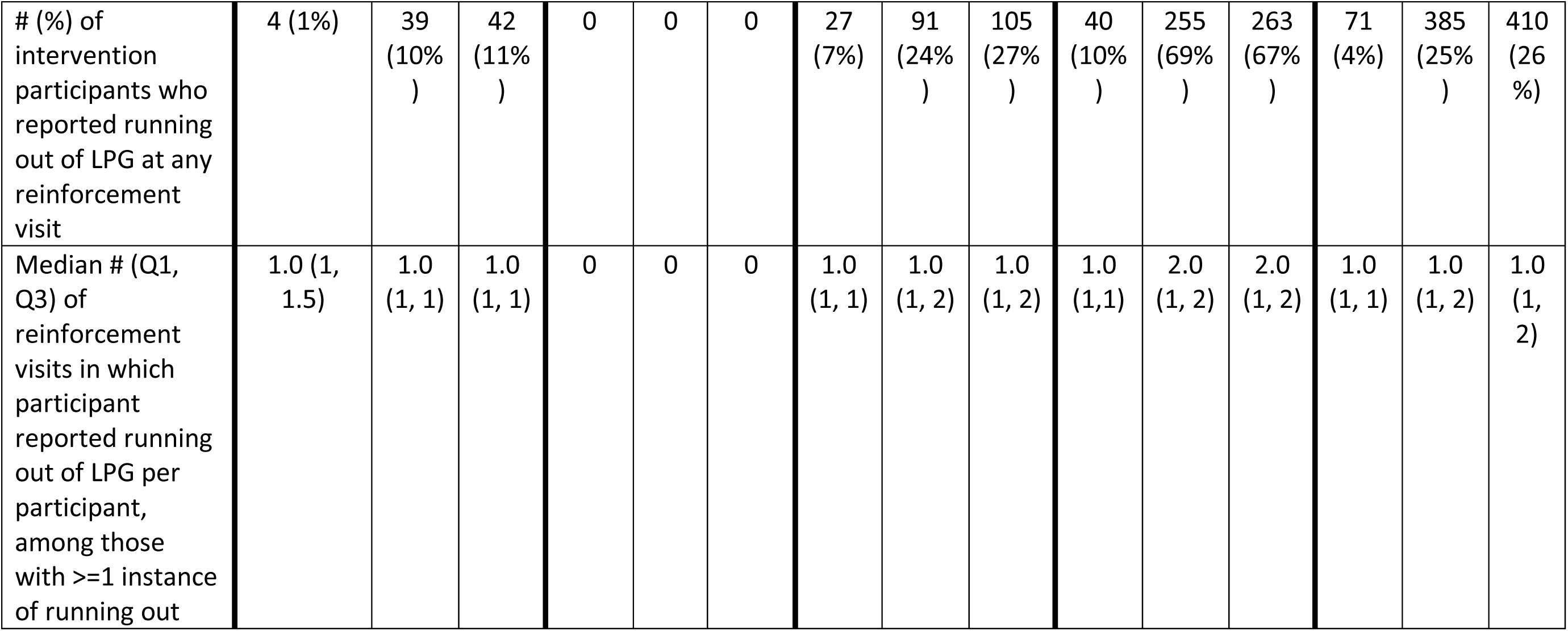
Reports of general problems or concerns with the LPG stove and running out of LPG at behavioral reinforcement visits in pregnancy (“preg”), the post-birth or infancy period (“infcy”), and total across the full trial (“full”) among intervention participants.

|  | Guatemala |  |  | India |  |  | Peru |  |  | Rwanda |  |  | Total |  |  |
| --- | --- | --- | --- | --- | --- | --- | --- | --- | --- | --- | --- | --- | --- | --- | --- |
| Study period in relation to the baby’s birth | Preg | Infcy | Full | Preg | Infcy | Full | Preg | Infcy | Full | Preg | Infcy | Full | Preg | Infcy | Full |
| N Intervention Participants | 400 | 382 | 400 | 398 | 384 | 398 | 394 | 379 | 394 | 392 | 372 | 392 | 1584 | 1517 | 1584 |
| # (%) of intervention participants who indicated a problem or concern about LPG use at any reinforcement visit | 58 (15%) | 145 (38%) | 169 (42%) | 3 (1%) | 0 | 3 (1%) | 146 (37%) | 140 (37%) | 211 (54%) | 68 (17%) | 291 (78%) | 302 (77%) | 275 (17%) | 576 (38%) | 685 (43%) |
| Median (Q1, Q3) # of reinforcement visits in which participant indicated a problem or concern per participant, among those with >=1 problems/ concerns | 1 (1, 1) | 1 (1, 2) | 1 (1, 2) | 1 (1, 1) | 0 | 1 (1, 1) | 1 (1, 2) | 2 (1, 3) | 2 (1, 3) | 1 (1, 1) | 2 (1, 3) | 2 (1, 3) | 1 (1, 2) | 2 (1, 3) | 2 (1, 3) |
| # (%) of intervention participants who reported running out of LPG at any reinforcement visit | 4 (1%) | 39 (10%) | 42 (11%) | 0 | 0 | 0 | 27 (7%) | 91 (24%) | 105 (27%) | 40 (10%) | 255 (69%) | 263 (67%) | 71 (4%) | 385 (25%) | 410 (26%) |
| Median # (Q1, Q3) of reinforcement visits in which participant reported running out of LPG per participant, among those with >=1 instance of running out | 1.0 (1, 1.5) | 1.0 (1, 1) | 1.0 (1, 1) | 0 | 0 | 0 | 1.0 (1, 1) | 1.0 (1, 2) | 1.0 (1, 2) | 1.0 (1,1) | 2.0 (1, 2) | 2.0 (1, 2) | 1.0 (1, 1) | 1.0 (1, 2) | 1.0 (1, 2) |

**Table S2.** Types of repairs made to LPG equipment by country and overall.

|  | Guatemala | India | Peru | Rwanda | Total |
| --- | --- | --- | --- | --- | --- |
| Total repair visits made | 348 | 50 | 547 | 641 | 1586 |
| Types of repairs |  |  |  |  |  |
| Stove* | 208 (60%) | 32 (64%) | 443 (81%) | 611 (95%) | 1294 (82%) |
| Knobs/burners | 23 (7%) | 23 (46%) | 309 (56%) | 513 (80%) | 868 (55%) |
| Stove valves | 187 (54%) | 7 (14%) | 113 (21%) | 4 (1%) | 311 (20%) |
| Stove or part replaced | 1 (0.3%) | 3 (6%) | 41 (8%) | 90 (14%) | 135 (9%) |
| LPG cylinder/regulator | 3 (1%) | 13 (26%) | 54 (10%) | 13 (2%) | 83 (5%) |
| Hose and connectors | 1 (0.3%) | 7 (14%) | 127 (23%) | 8 (1%) | 143 (9%) |
| Switch valve | 139 (40%) | N/A | N/A | 12 (2%) | 151 (15%) |

\*Only includes most commonly reported stove problems; a few “other” responses not summarized here
NOTE: Numbers may add up to greater than 100% because multiple problems may have been fixed at the same visit.

**Table S3.**
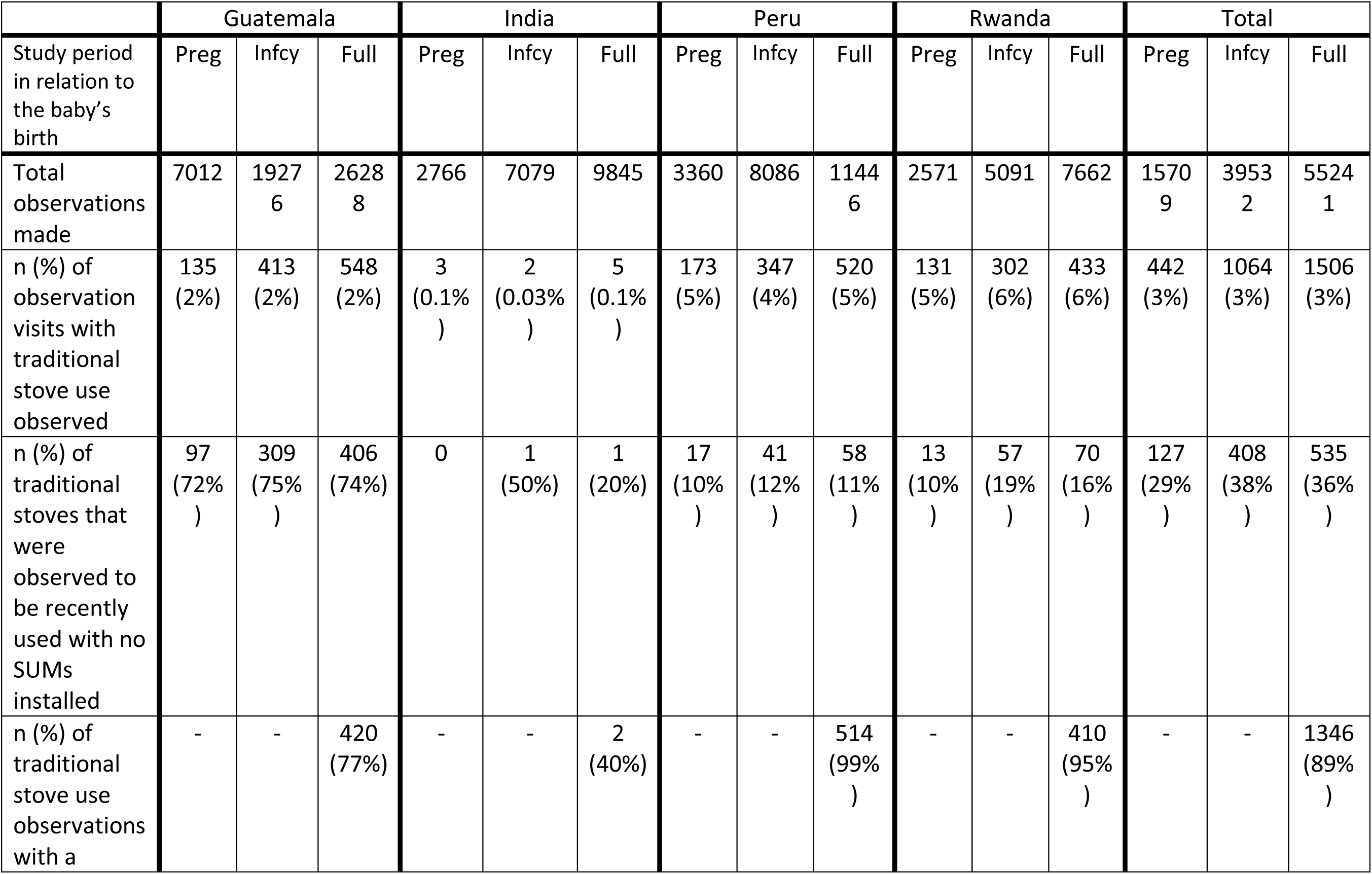
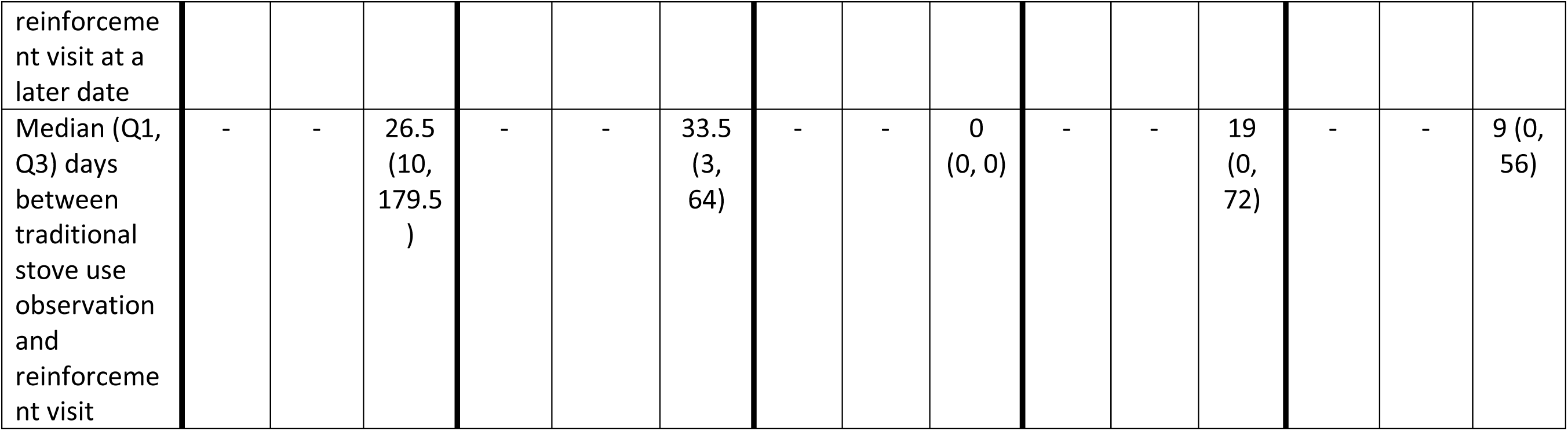
Observations of traditional stove use and follow-up behavioral reinforcement in pregnancy (“preg”), the post-birth or infancy period (“infcy”), and total across the full trial (“full”) among intervention participants.

**Table S4.** Stove use monitoring in intervention households by country.

|  | Guatemala | India | Peru | Rwanda | Total |
| --- | --- | --- | --- | --- | --- |
| **# Intervention households who received LPG stove** | **400** | **398** | **394** | **392** | **1584** |
| # Households who voluntarily removed TS at LPG delivery and did not re-install it during pregnancy (%) | 265 (66%) | 214 (54%) | 8 (2%) | 10 (3%) | 497 (31%) |
| # Households missing SUMs data during pregnancy for unknown reason* (%) | 2 (0.5%) | 2 (0.5%) | 1 (0.3%) | 43 (11%) | 48 (3%) |
| **Total households with valid SUMs during pregnancy** | **133** | **182** | **385** | **339** | **1039** |
| # Households with miscarriage or stillbirth | 16 | 10 | 9 | 13 | 48 |
| # Households with infant death or drop-out <14 days after birth | 2 | 4 | 6 | 7 | 19 |
| **# live births with post-birth follow-up of mother >14 days** | **382** | **384** | **379** | **372** | **1517** |
| # Households who removed TS at LPG delivery and did not re-install it during pregnancy or post-birth (%) | 222 (58%) | 204 (53%) | 5 (1%) | 5 (1%) | 436 (29%) |
| # Households who removed TS during pregnancy or <14 days after birth and did not re-install post-birth (%) | 0 | 28 (7%) | 33 (9%) | 0 | 61 (4%) |
| # Households missing SUMs data post-birth for unknown reason* (%) | 58 (15%) | 13 (3%) | 37 (10%) | 35 (9%) | 143 (9%) |
| **Total households with valid SUMs post-birth** | **102** | **139** | **304** | **332** | **877** |
| **Total households with valid SUMs in pregnancy and/or post-birth periods** | **164** | **184** | **387** | **360** | **1095** |

TS=Traditional stove; SUMs=Stove use monitors; LPG=Liquefied petroleum gas
\*Reasons for missing SUMs data may include SUM device errors, participant manipulation or removal of the SUMs, participant misclassification (i.e., failure to record removal of a traditional stove, or mistakenly indicating that participant retained a traditional stove when they had removed it), fieldworker errors in SUM installation, inability to match SUMs data to a household, among others.

**Table S5.** Additional details on traditional stove monitoring and traditional stove use (TSU) based on SUMs data from intervention households in pregnancy (“preg”), the post-birth or infancy period (“infcy”), and total across the full trial (“full”). This table complements the data in Table 6 of the main paper.

|  | Guatemala |  |  | India |  |  | Peru |  |  | Rwanda |  |  | Total |  |  |
| --- | --- | --- | --- | --- | --- | --- | --- | --- | --- | --- | --- | --- | --- | --- | --- |
| Study period in relation to the baby's birth | Preg | Infcy | Full | Preg | Infcy | Full | Preg | Infcy | Full | Preg | Infcy | Full | Preg | Infcy | Full |
| Total households with valid SUMs data | 133 | 102 | 164 | 182 | 139 | 184 | 385 | 304 | 387 | 339 | 332 | 360 | 1039 | 877 | 1095 |
| Days with stove-use-monitoring per household: median (Q1, Q3) | 99<br>(29, 146) | 358<br>(153.2, 392.8) | 169.5<br>(41, 501.2) | 127<br>(91.2, 149.5) | 366<br>(315.5, 376) | 460<br>(156.5, 510) | 145<br>(121, 170) | 370<br>(205.2, 384) | 471<br>(196.5, 538) | 134<br>(95, 161) | 338<br>(243.8, 387) | 451.5<br>(327.8, 511.2) | 134<br>(97.5, 161) | 363<br>(234, 385) | 435<br>(193, 521) |
| Percent of monitored days with TSU detected: mean (range) | 3.8<br>(0 - 95.2) | 6.8<br>(0 - 100) | 4.6<br>(0 - 94) | 1.5<br>(0 - 53.2) | 0.3<br>(0 - 6.1) | 1.1<br>(0 - 53.2) | 4.3<br>(0 - 81) | 3.6<br>(0 - 51.8) | 4.3<br>(0 - 81) | 1.8<br>(0 - 94.1) | 2.1<br>(0 - 41.6) | 1.9<br>(0 - 31.5) | 2.9<br>(0 - 95.2) | 2.9<br>(0 - 100) | 3.0<br>(0 - 94) |
| Households with no SUM-detected TSU: N (%) | 105<br>(78.9 %) | 44<br>(43.1 %) | 96<br>(58.5 %) | 143<br>(78.6 %) | 113<br>(81.3 %) | 129<br>(70.1 %) | 177<br>(46%) | 145<br>(47.7 %) | 126<br>(32.6%) | 194<br>(57.2 %) | 114<br>(34.3 %) | 106<br>(29.4 %) | 619<br>(59.6 %) | 416<br>(47.4 %) | 457<br>(41.7%) |
| Household<br>s with < 1<br>day with<br>TSU per 30<br>days of<br>monitoring | 117<br>(88.0<br>%) | 72<br>(70.6<br>%) | 133<br>(81.1<br>%) | 167<br>(91.8<br>%) | 135<br>(97.1<br>%) | 172<br>(93.5<br>%) | 288<br>(74.8<br>%) | 228<br>(75%) | 273<br>(70.5%) | 293<br>(86.4<br>%) | 274<br>(82.5<br>%) | 307<br>(85.3<br>%) | 865<br>(83.3<br>%) | 709<br>(80.8<br>%) | 885<br>(80.8%) |

**Table S6.**
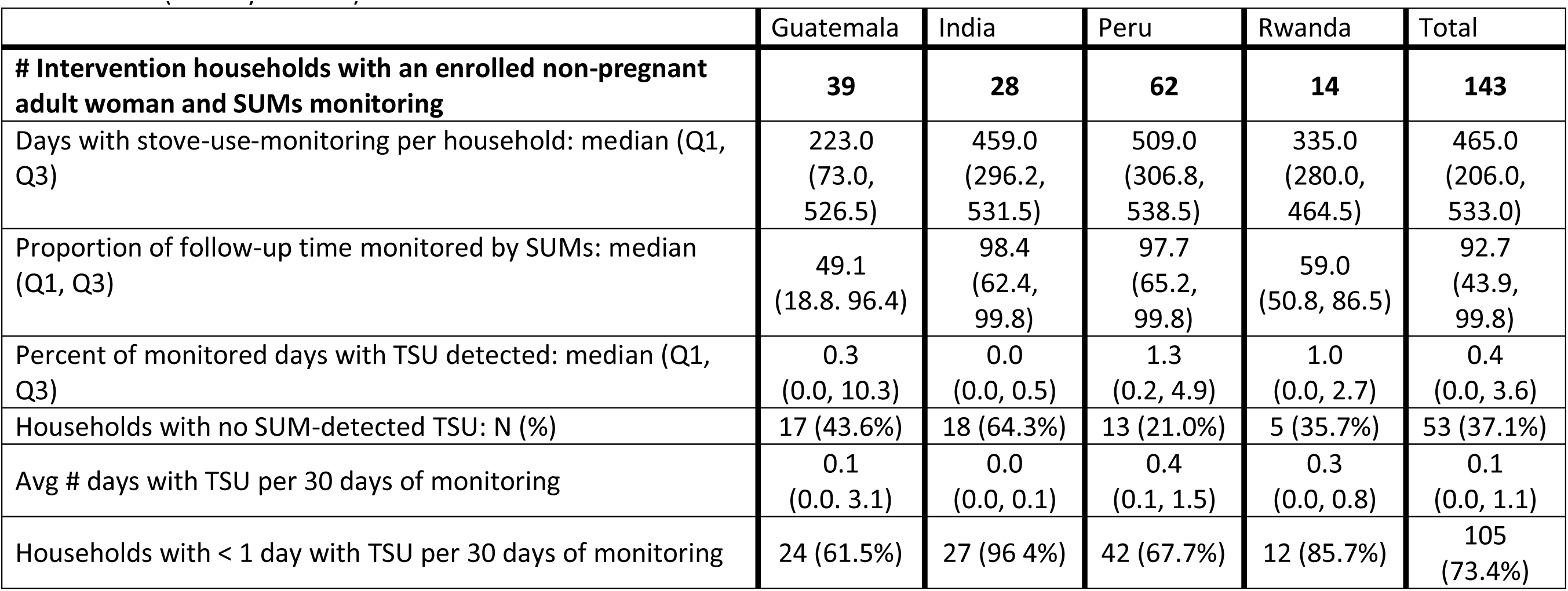
Traditional stove use (TSU) based on SUMs data from the subset of intervention households with an enrolled non-pregnant adult woman (40-79 years old) across the full trial.

|  | Guatemala | India | Peru | Rwanda | Total |
| --- | --- | --- | --- | --- | --- |
| **# Intervention households with an enrolled non-pregnant adult woman and SUMs monitoring** | **39** | **28** | **62** | **14** | **143** |
| Days with stove-use-monitoring per household: median (Q1, Q3) | 223.0<br>(73.0, 526.5) | 459.0<br>(296.2, 531.5) | 509.0<br>(306.8, 538.5) | 335.0<br>(280.0, 464.5) | 465.0<br>(206.0, 533.0) |
| Proportion of follow-up time monitored by SUMs: median (Q1, Q3) | 49.1<br>(18.8, 96.4) | 98.4<br>(62.4, 99.8) | 97.7<br>(65.2, 99.8) | 59.0<br>(50.8, 86.5) | 92.7<br>(43.9, 99.8) |
| Percent of monitored days with TSU detected: median (Q1, Q3) | 0.3<br>(0.0, 10.3) | 0.0<br>(0.0, 0.5) | 1.3<br>(0.2, 4.9) | 1.0<br>(0.0, 2.7) | 0.4<br>(0.0, 3.6) |
| Households with no SUM-detected TSU: N (%) | 17 (43.6%) | 18 (64.3%) | 13 (21.0%) | 5 (35.7%) | 53 (37.1%) |
| Avg # days with TSU per 30 days of monitoring | 0.1<br>(0.0, 3.1) | 0.0<br>(0.0, 0.1) | 0.4<br>(0.1, 1.5) | 0.3<br>(0.0, 0.8) | 0.1<br>(0.0, 1.1) |
| Households with < 1 day with TSU per 30 days of monitoring | 24 (61.5%) | 27 (96.4%) | 42 (67.7%) | 12 (85.7%) | 105<br>(73.4%) |

**Table S7.** Traditional stove use based on SUMs data from intervention households pre-COVID-19 and post-COVID-19.

|  | Guatemala |  | India |  | Peru |  | Rwanda |  | Total |  |
| --- | --- | --- | --- | --- | --- | --- | --- | --- | --- | --- |
|  | Pre-COVID | Post-COVID | Pre-COVID | Post-COVID | Pre-COVID | Post-COVID | Pre-COVID | Post-COVID | Pre-COVID | Post-COVID |
| Total households with valid SUMs data | 163 | 74 | 184 | 99 | 383 | 258 | 357 | 226 | 1087 | 657 |
| Days with stove-use-monitoring per household: median (Q1, Q3) | 89<br>(38, 302) | 132<br>(72.2, 220.5) | 247.5<br>(121, 349.2) | 196.0<br>(114.5, 263) | 181<br>(90, 300.5) | 218.5<br>(133.2, 354.8) | 292<br>(192, 399) | 162<br>(89.2, 239.5) | 225<br>(110.5, 350) | 188<br>(108, 279) |
| Proportion of follow-up time monitored by SUMs | 30.5<br>(11.9, 95.9) | 98.5<br>(94.2, 99.3) | 96.8<br>(44.7, 100) | 99.2<br>(97.5, 99.6) | 100<br>(72.5, 100) | 99.2<br>(65.9, 99.6) | 91.4<br>(71.7, 100) | 89.3<br>(65.3, 99.2) | 95.9<br>(52.4, 100) | 98.7<br>(72.9, 99.5) |
| Percent of monitored days with TSU detected: median (Q1, Q3) | 0.0<br>(0.0, 0.9) | 0.0<br>(0.0, 4.2) | 0.0<br>(0.0, 0.0) | 0.0<br>(0.0, 0.0) | 0.2<br>(0.0, 2.3) | 0.7<br>(0.0, 5.5) | 0.4<br>(0.0, 1.6) | 0.7<br>(0.0, 3.5) | 0.0<br>(0.0, 1.6) | 0.0<br>(0.0, 3.4) |
| Households with no SUM-detected TSU: N (%) | 106<br>(65%) | 39<br>(52.7%) | 140<br>(76.1%) | 82<br>(82.8%) | 190<br>(49.6%) | 110<br>(42.6%) | 144<br>(40.3%) | 98<br>(43.4%) | 580<br>(53.4%) | 329<br>(50.1%) |
| Avg # days with TSU per 30 days of monitoring | 0.0<br>(0.0, 0.3) | 0.0<br>(0.0, 1.3) | 0.0<br>(0.0, 0.0) | 0.0<br>(0.0, 0.0) | 0.1<br>(0.0, 0.7) | 0.2<br>(0.0, 1.6) | 0.1<br>(0.0, 0.5) | 0.2<br>(0.0, 1.1) | 0.0<br>(0.0, 0.5) | 0.0<br>(0.0, 1.0) |
| Households with < 1 day with TSU per 30 days of monitoring | 135<br>(82.8%) | 52<br>(70.3%) | 171<br>(92.9%) | 98<br>(99%) | 303<br>(79.1%) | 175<br>(67.8%) | 315<br>(88.2%) | 164<br>(72.6%) | 924<br>(85%) | 489<br>(74.4%) |

**Table S8.**
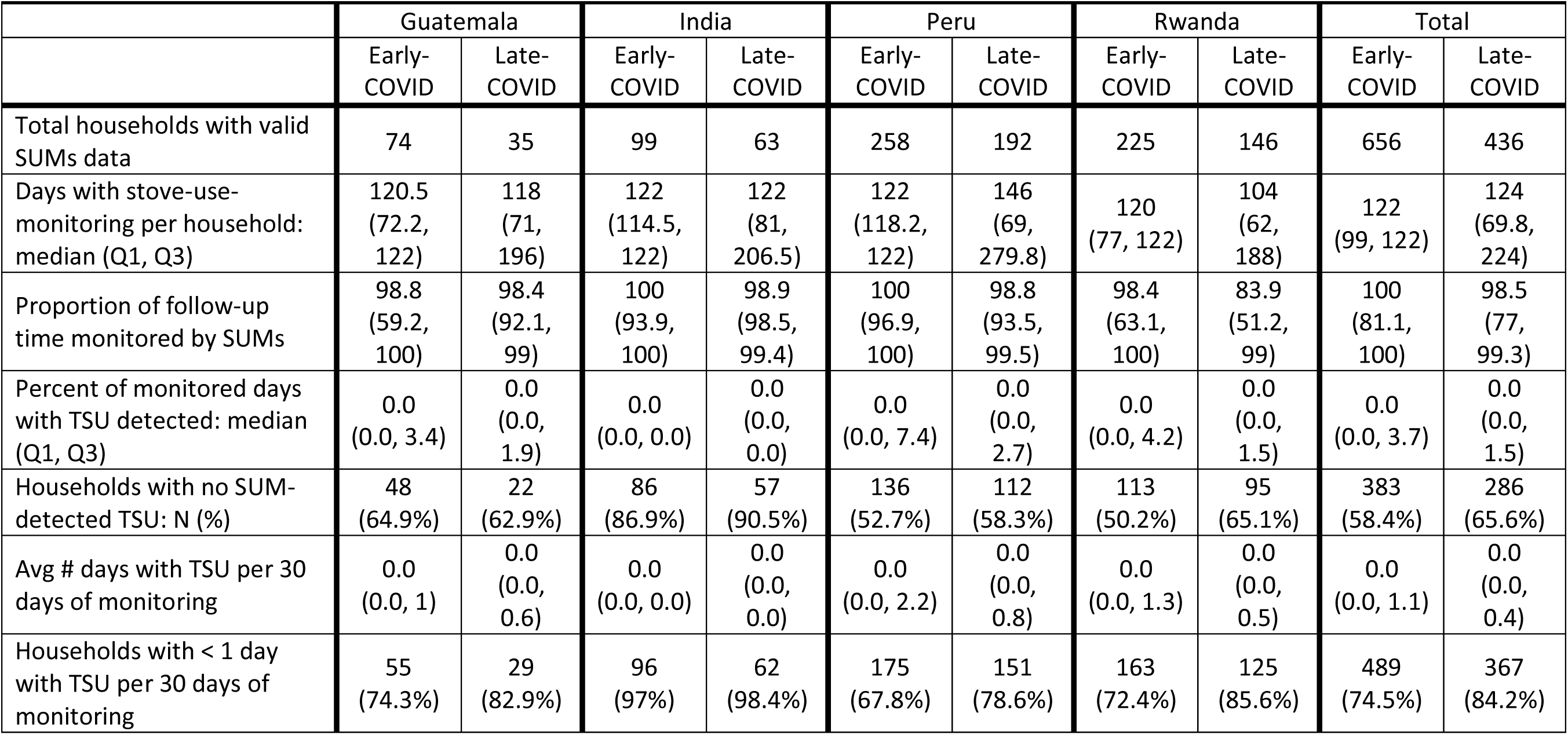
Traditional stove use based on SUMs data from intervention households in early-COVID-19 (March 17, 2020 – July 17, 2020) and late-COVID-19 (after July 17, 2020) periods.

|  | Guatemala |  | India |  | Peru |  | Rwanda |  | Total |  |
| --- | --- | --- | --- | --- | --- | --- | --- | --- | --- | --- |
|  | Early-COVID | Late-COVID | Early-COVID | Late-COVID | Early-COVID | Late-COVID | Early-COVID | Late-COVID | Early-COVID | Late-COVID |
| Total households with valid SUMs data | 74 | 35 | 99 | 63 | 258 | 192 | 225 | 146 | 656 | 436 |
| Days with stove-use-monitoring per household: median (Q1, Q3) | 120.5<br>(72.2, 122) | 118<br>(71, 196) | 122<br>(114.5, 122) | 122<br>(81, 206.5) | 122<br>(118.2, 122) | 146<br>(69, 279.8) | 120<br>(77, 122) | 104<br>(62, 188) | 122<br>(99, 122) | 124<br>(69.8, 224) |
| Proportion of follow-up time monitored by SUMs | 98.8<br>(59.2, 100) | 98.4<br>(92.1, 99) | 100<br>(93.9, 100) | 98.9<br>(98.5, 99.4) | 100<br>(96.9, 100) | 98.8<br>(93.5, 99.5) | 98.4<br>(63.1, 100) | 83.9<br>(51.2, 99) | 100<br>(81.1, 100) | 98.5<br>(77, 99.3) |
| Percent of monitored days with TSU detected: median (Q1, Q3) | 0.0<br>(0.0, 3.4) | 0.0<br>(0.0, 1.9) | 0.0<br>(0.0, 0.0) | 0.0<br>(0.0, 0.0) | 0.0<br>(0.0, 7.4) | 0.0<br>(0.0, 2.7) | 0.0<br>(0.0, 4.2) | 0.0<br>(0.0, 1.5) | 0.0<br>(0.0, 3.7) | 0.0<br>(0.0, 1.5) |
| Households with no SUM-detected TSU: N (%) | 48<br>(64.9%) | 22<br>(62.9%) | 86<br>(86.9%) | 57<br>(90.5%) | 136<br>(52.7%) | 112<br>(58.3%) | 113<br>(50.2%) | 95<br>(65.1%) | 383<br>(58.4%) | 286<br>(65.6%) |
| Avg # days with TSU per 30 days of monitoring | 0.0<br>(0.0, 1) | 0.0<br>(0.0, 0.6) | 0.0<br>(0.0, 0.0) | 0.0<br>(0.0, 0.0) | 0.0<br>(0.0, 2.2) | 0.0<br>(0.0, 0.8) | 0.0<br>(0.0, 1.3) | 0.0<br>(0.0, 0.5) | 0.0<br>(0.0, 1.1) | 0.0<br>(0.0, 0.4) |
| Households with < 1 day with TSU per 30 days of monitoring | 55<br>(74.3%) | 29<br>(82.9%) | 96<br>(97%) | 62<br>(98.4%) | 175<br>(67.8%) | 151<br>(78.6%) | 163<br>(72.4%) | 125<br>(85.6%) | 489<br>(74.5%) | 367<br>(84.2%) |

**Table S9.**
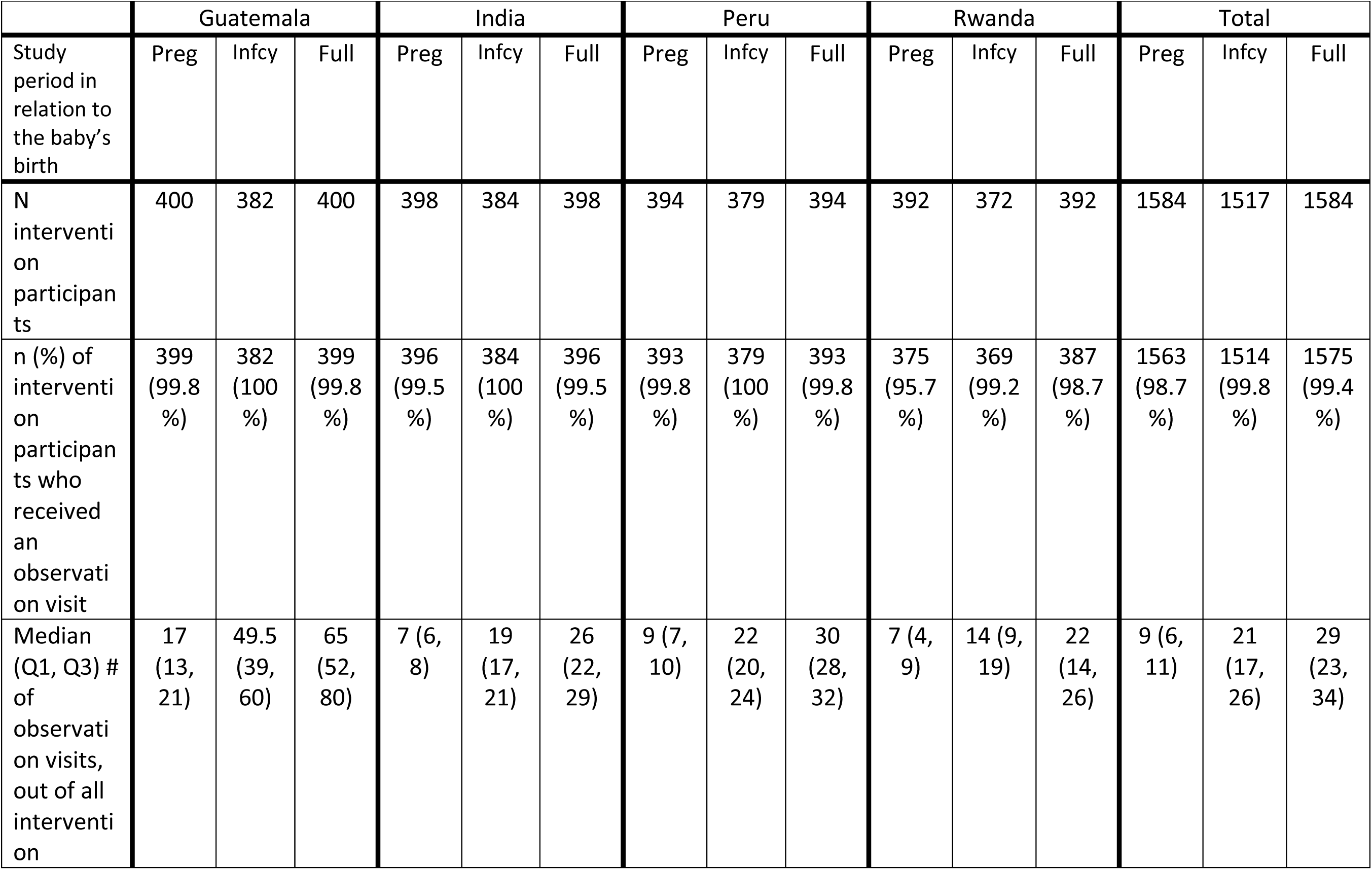
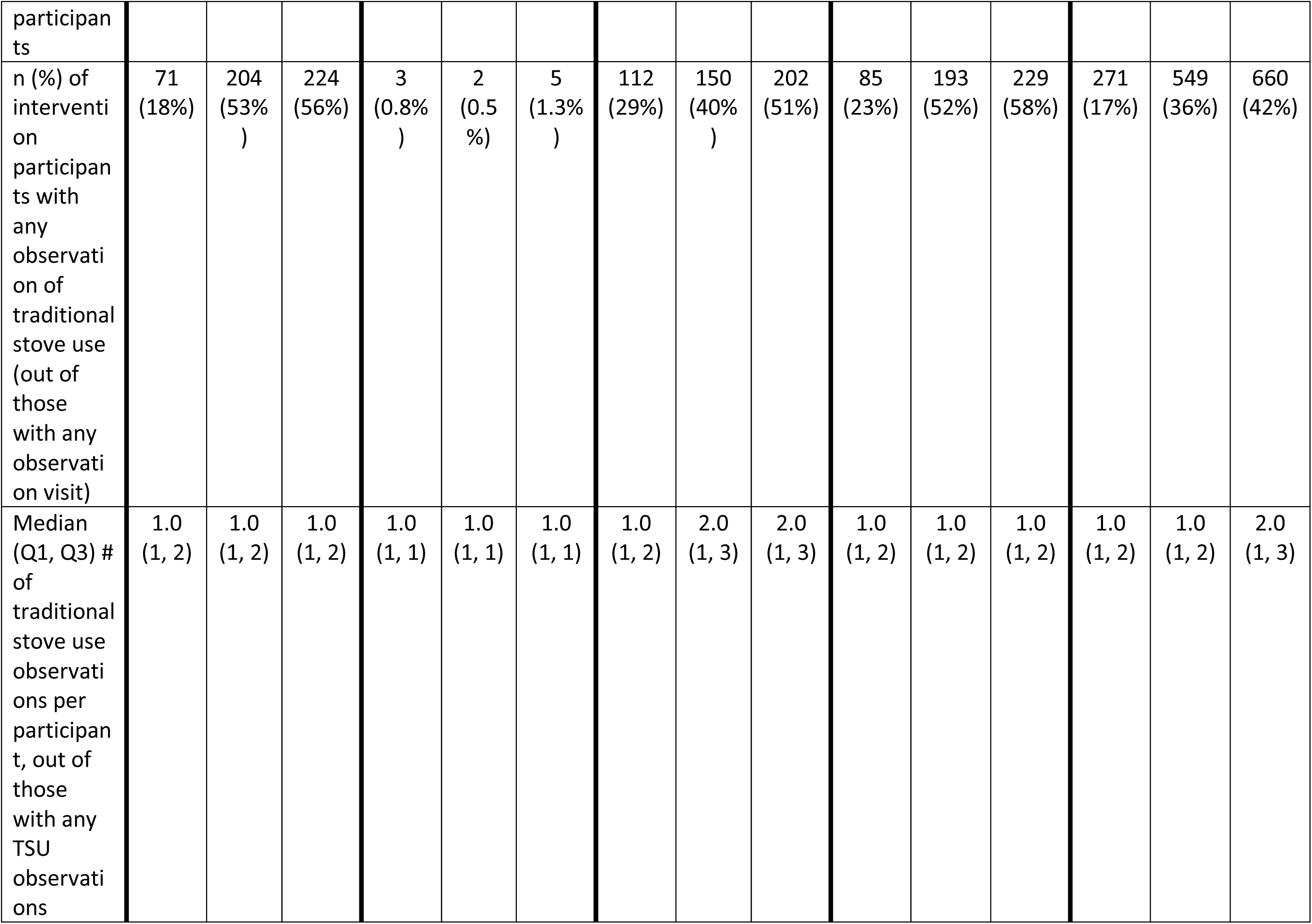
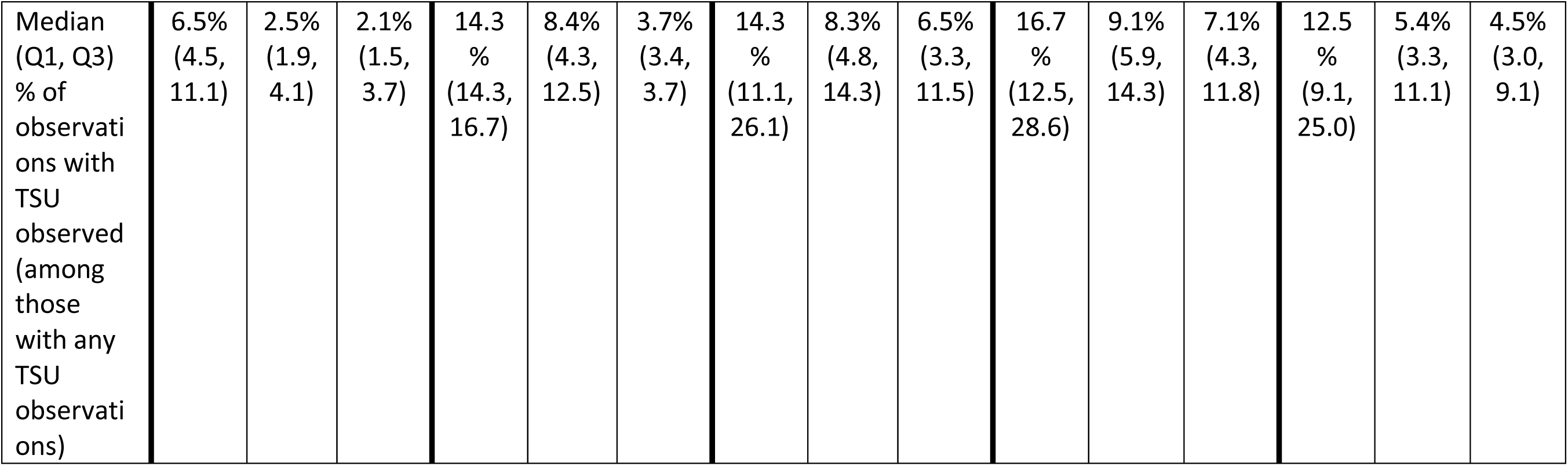
Percent of intervention participants with observations of traditional stove use during pregnancy (“preg”), post-birth or infancy (“infcy”), and total across the full trial (“full”), and extent to which traditional stove use was observed per participant.

**Table S10.**
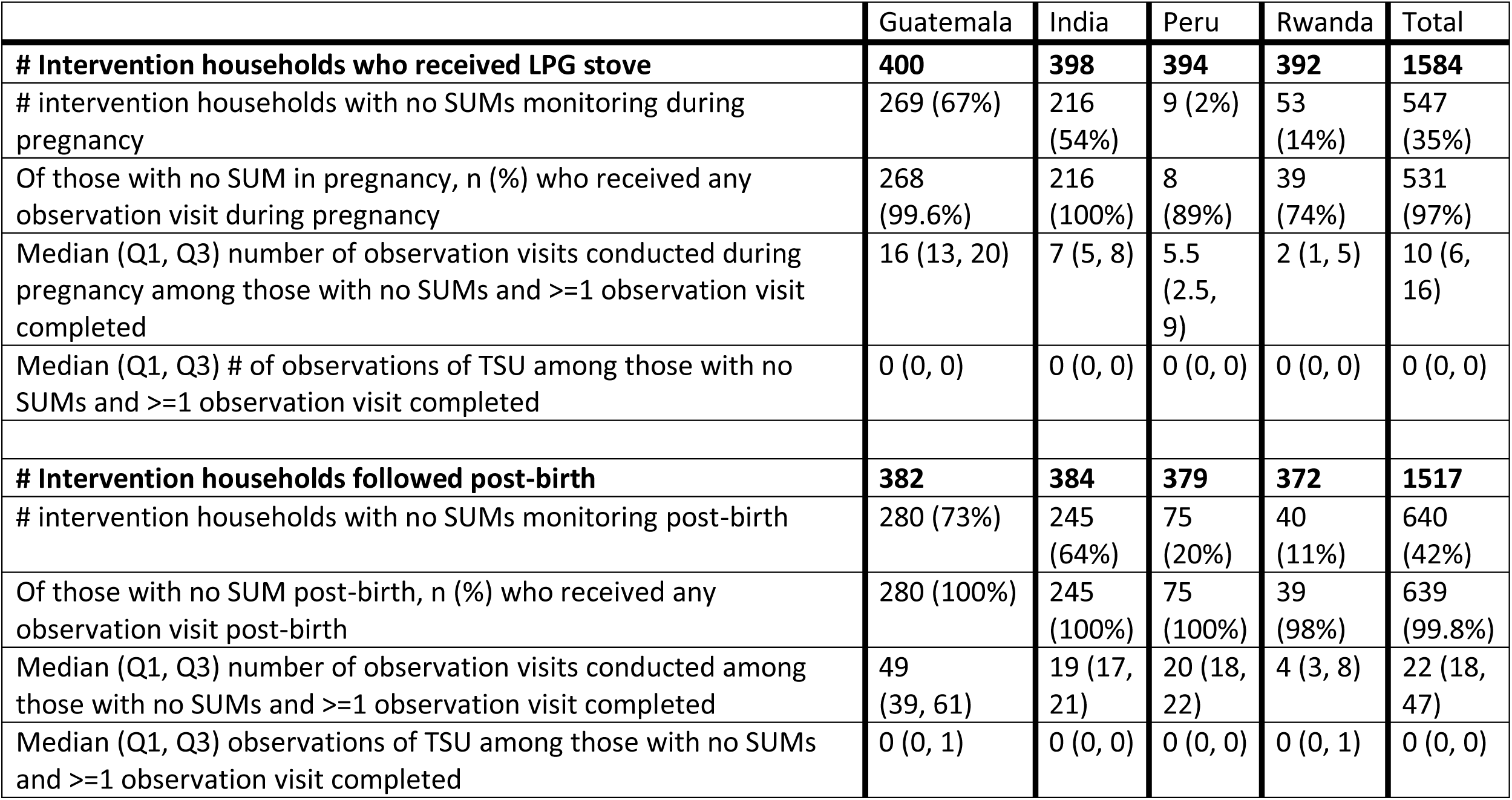
Observations of traditional stove use in intervention households missing SUMs data.

**Table S11.** Percent of intervention and control participants who ever moved during the trial, including use of biomass by intervention participants during moves and use of clean fuels by control participants during moves.

|  | Guatemala |  | India |  | Peru |  | Rwanda |  | Total |  |
| --- | --- | --- | --- | --- | --- | --- | --- | --- | --- | --- |
|  | Intvn | Cntrl | Intvn | Cntrl | Intvn | Cntrl | Intvn | Cntrl | Intvn | Cntrl |
| n | 400 | 400 | 398 | 399 | 394 | 402 | 392 | 404 | 1584 | 1605 |
| # (%) households that ever moved | 65<br>(16%) | 77<br>(19%) | 96<br>(24%) | 98<br>(25%) | 116<br>(29%) | 131<br>(33%) | 71<br>(18%) | 60<br>(15%) | 348<br>(22%) | 366<br>(23%) |
| # (%) Intervention participants who moved to house where biomass is used at all | 13<br>(3%) | --- | 52<br>(13%) | --- | 10<br>(3%) | --- | 4<br>(1%) | --- | 79<br>(5%) | --- |
| Median (Q1, Q3) days spent by intervention participants in house where biomass is used at all | 104<br>(83, 121) | --- | 153<br>(86.5, 223.5) | --- | 86.5<br>(26, 168) | --- | 94<br>(64, 122) | --- | 136<br>(71, 215) | --- |
| # (%) Control participants who moved to house where clean fuel is used exclusively | --- | 2<br>(0.5%) | --- | 7<br>(2%) | --- | 42<br>(10%) | --- | 0 | --- | 51<br>(3%) |
| Median (Q1, Q3) days spent by control participants in house where clean fuel is used exclusively | --- | 506<br>(471, 540) | --- | 147<br>(18, 309) | --- | 136<br>(84, 220) | --- | 0 | --- | 141<br>(79, 246) |

The most frequent reasons for moves included visiting the mother's or mother-in-law's house, moving to a new personal residence, and moving to a seasonal residence. Visiting the mother's or mother-in-law's house was the most common reason for moves in Guatemala, India, and Peru, while moving to a new personal residence was most common in Rwanda. Reasons were similar between intervention and control participants. Across countries, control participants who moved to a house where clean fuel was used exclusively spent a median (Q1, Q3) of 141 (79, 246) days in the new or temporary home. Intervention participants who moved to a home where biomass is used at all spent a median (Q1, Q3) of 136 (71, 215) days in the new or temporary home, but only 29% (n=23) reported being the primary cook in that home.

## REFERENCES

1. IEA, IRENA, UNSD, World Bank, WHO. Tracking SDG 7: The energy progress report. World Bank, Washington DC. 2022.

2. Fandiño-Del-Rio Magdalena, Kephart Josiah L, Williams Kendra N, et al. Household air pollution concentrations after liquefied petroleum gas interventions in rural Peru: Findings from a one-year randomized controlled trial followed by a one-year pragmatic crossover trial. Environ Health Perspect.; 130(5):057007. doi: 10.1289/EHP10054.

3. Chillrud SN, Ae-Ngibise K, Gould CF, et al. The effect of clean cooking interventions on mother and child personal exposure to air pollution: Results from the Ghana randomized air pollution and health study (GRAPHS). Journal of Exposure Science & Environmental Epidemiology. 2021.

4. GBD 2019 Risk Factors Collaborators. Global burden of 87 risk factors in 204 countries and territories, 1990-2019: A systematic analysis for the global burden of disease study 2019. Lancet. 2020;396(10258):1223–1249.

5. Lee KK, Bing R, Kiang J, et al. Adverse health effects associated with household air pollution: A systematic review, meta-analysis, and burden estimation study. The Lancet Global Health. 2020;8(11):e1427–e1434. doi: 10.1016/S2214-109X(20)30343-0.

6. Balakrishnan K, Steenland K, Clasen T, et al. Exposure–response relationships for personal exposure to fine particulate matter (PM_2·5_), carbon monoxide, and black carbon and birthweight: Results from the multi-country household air pollution intervention network (HAPIN) trial. MedRxiv. 2022:2022.08.06.22278373. doi: 10.1101/2022.08.06.22278373.

7. Smith KR, McCracken JP, Weber MW, et al. Effect of reduction in household air pollution on childhood pneumonia in Guatemala (RESPIRE): A randomised controlled trial. Lancet. 2011;378(9804):1717–1726.

8. Katz J, Tielsch JM, Khatry SK, et al. Impact of improved biomass and liquid petroleum gas stoves on birth outcomes in rural Nepal: Results of 2 randomized trials. Glob Health Sci Pract. 2020;8(3):372–382. doi: 10.9745/GHSP-D-20-00011.

9. Jack DW, Ae-Ngibise KA, Gould CF, et al. A cluster randomised trial of cookstove interventions to improve infant health in Ghana. BMJ Glob Health. 2021;6(8):e005599. doi: 10.1136/bmjgh-005599.

10. Alexander DA, Northcross A, Karrison T, et al. Pregnancy outcomes and ethanol cook stove intervention: A randomized-controlled trial in Ibadan, Nigeria. Environ Int. 2018;111:152–163.

11. Clasen TF, Chang HH, Thompson LM, et al. Liquefied petroleum gas or biomass for cooking and effects on birth weight. N Engl J Med. 2022. doi: 10.1056/NEJMoa2206734.

12. Bruce N, Pope D, Rehfuess E, Balakrishnan K, Adair-Rohani H, Dora C. WHO indoor air quality guidelines on household fuel combustion: Strategy implications of new evidence on interventions and exposure-risk functions. Atmos Environ. 2015;106:451–457.

13. Checkley W, Williams KN, Kephart JL, et al. Effects of a household air pollution intervention with liquefied petroleum gas on cardiopulmonary outcomes in Peru. A randomized controlled trial. Am J Respir Crit Care Med. 2021;203(11):1386–1397. doi: 10.1164/rccm.202006-2319OC.

14. Clasen T, Checkley W, Peel JL, et al. Design and rationale of the HAPIN study: A multicountry randomized controlled trial to assess the effect of liquefied petroleum gas stove and continuous fuel distribution. Environ Health Perspect. 2020;128(4):047008.

15. Johnson MA, Chiang RA. Quantitative guidance for stove usage and performance to achieve health and environmental targets. Environ Health Perspect. 2015;123(8):820–826.

16. Williams KN, Thompson LM, Sakas Z, et al. Designing a comprehensive behaviour change intervention to promote and monitor exclusive use of liquefied petroleum gas stoves for the household air pollution intervention network (HAPIN) trial. BMJ Open. 2020;10(9).

17. Quinn AK, Williams KN, Thompson LM, et al. Fidelity and adherence to a liquefied petroleum gas stove and fuel intervention during gestation: The multi-country household air pollution intervention network (HAPIN) randomized controlled trial. International Journal of Environmental Research and Public Health. 2021;18(23). doi: 10.3390/ijerph182312592.

18. Carrión D, Prah R, Gould CF, et al. Using longitudinal survey and sensor data to understand the social and ecological determinants of clean fuels use and discontinuance in rural ghana. Environmental Research Communications. 2020;2(9):095003. doi: 10.1088/2515-7620/abb831.

19. Aung TW, Baumgartner J, Jain G, et al. Effect on blood pressure and eye health symptoms in a climate-financed randomized cookstove intervention study in rural India. Environ Res. 2018;166:658–667. doi: 10.1016/j.envres.2018.06.044.

20. Gill-Wiehl A, Brown T, Smith K. The need to prioritize consumption: A difference-in-differences approach to analyze the total effect of India’s below-the-poverty-line policies on LPG use. Energy Policy. 2022;164:112915. doi: 10.1016/j.enpol.2022.112915.

21. Ravindra K, Kaur-Sidhu M, Mor S, Chakma J, Pillarisetti A. Impact of the COVID-19 pandemic on clean fuel programmes in India and ensuring sustainability for household energy needs. Environ Int. 2021;147:106335. doi: 10.1016/j.envint.2020.106335.

22. Williams KN, Kephart JL, Fandino-Del-Rio M, et al. Beyond cost: Exploring fuel choices and the socio-cultural dynamics of liquefied petroleum gas stove adoption in Peru. Energy Research & Social Science. 2020;66:101591.

23. Puzzolo E, Pope D, Stanistreet D, Rehfuess EA, Bruce NG. Clean fuels for resource-poor settings: A systematic review of barriers and enablers to adoption and sustained use. Environ Res. 2016;146:218–234.

24. Liao J, Kirby MA, Pillarisetti A, et al. LPG stove and fuel intervention among pregnant women reduce fine particle air pollution exposures in three countries: Pilot results from the HAPIN trial. Environmental Pollution. 2021;291:118198. doi: 10.1016/j.envpol.2021.118198.

25. Gould CF, Jagoe K, Moreno AI, et al. Prevalent degradation and patterns of use, maintenance, repair, and access to post-acquisition services for biomass stoves in Peru. Energy Sustain Dev. 2018;45:79.

26. Wilson DL, Williams KN, Pillarisetti A. An integrated sensor data logging, survey, and analytics platform for field research and its application in HAPIN, a multi-center household energy intervention trial. Sustainability. 2020;12(5).

27. Simkovich SM, Thompson LM, Clark ML, et al. A risk assessment tool for resumption of research activities during the COVID-19 pandemic for field trials in low resource settings. BMC Medical Research Methodology. 2021;21(1):68. doi: 10.1186/s12874-021-01232-x.

